# Exploring the role of binge eating in the association between ADHD and BMI: A twin study

**DOI:** 10.64898/2026.05.28.26354354

**Authors:** Yuan You, Tom A. McAdams, Olakunle Oginni, Chaoyu Liu, Moritz Herle, Helena M.S. Zavos

**Affiliations:** Social, Genetic and Developmental Psychiatry Centre, Institute of Psychiatry, Psychology & Neuroscience, King’s College London, London, UK; Wolfson Centre for Young People’s Mental Health, Cardiff University, Cardiff, UK; Department of Psychiatry, Yale University, New Haven, USA; Department of Psychology, Institute of Psychiatry, Psychology & Neuroscience, King’s College London, UK

**Keywords:** ADHD, Binge eating, BMI, Twin study

## Abstract

**Objective:** ADHD has been associated with obesity indicators, including BMI, across the lifespan. A possible mechanism linking ADHD and BMI is binge eating. Previous research has found associations between ADHD, binge eating and BMI. However, the role of genetic and environmental influences on these associations remains unclear.

**Method:** We utilized data from the Twins Early Development Study (TEDS), comprising 3,675 monozygotic and 7,063 dizygotic twin pairs. ADHD symptoms in childhood and adolescence were assessed using parent-reported questionnaires. Adult ADHD symptoms were measured using both self-report and parent-report questionnaires. Phenotypic mediation models examined whether binge eating mediated the association between ADHD and BMI, without controlling for genetic confounding. Subsequently, the etiological architecture underlying the associations among the three traits across childhood, adolescence, and adulthood were investigated by incorporating genetic and environmental influences into the models.

**Results:** Binge eating significantly mediated the association between ADHD symptoms and BMI in both adolescence and adulthood. However, these mediation effects were no longer present once genetic and environmental influences were incorporated into the models. The best-fitting model in childhood, adolescence and adulthood was Cholesky decomposition models, where covariance between traits was explained by shared aetiology.

**Conclusions:** This twin study reveals shared liability across ADHD, binge eating, and BMI. The mediating role of binge eating in the relationship between ADHD symptoms and BMI was largely confounded by shared genetic influences. Intervention strategies could focus more on common underlying behavioural and self-regulatory mechanisms across these traits, as well as placing more emphasis on symptom patterns within families.

## Introduction

ADHD is characterized by symptoms of inattention, impulsivity, and hyperactivity (DSM-5; APA, 2013), and has been associated with obesity indicators, including BMI, across the lifespan (Cortese et al., 2016; Liu et al., 2021, Nigg et al., 2016). A possible mechanism linking ADHD and BMI is binge eating. Binge eating symptoms have been commonly reported in both individuals with ADHD and obesity (Cortese, Bernardina, & Mouren, 2007; de Zwaan et al., 2001; Svedlund, Norring, Ginsberg, & von Hausswolff-Juhlin, 2017). Understanding the mechanisms underlying the links between ADHD, binge eating behaviors and BMI is a priority both for people with lived experience of neurodiversity and for public health (Cortese et al., 2007; Kaisari, Dourish, & Higgs, 2017; Keller, Herle, Mandy, & Leno, 2024).

The association between ADHD and BMI has been well-established (Cortese et al., 2016). Phenotypic studies have shown that ADHD and its symptoms are associated with higher BMI and an increased risk of overweight from childhood through early adulthood (Fliers et al., 2013; Fuemmeler, Østbye, Yang, McClernon, & Kollins, 2011). Mendelian randomization analyses using polygenic scores have indicated bidirectional causal effects between ADHD and BMI, with genetic liability to ADHD influencing BMI and, conversely, genetic liability to BMI influencing ADHD (Liu et al., 2021).

Binge eating has been proposed as a major risk factor for obesity among individuals with ADHD (Cortese et al., 2007; Svedlund et al., 2017). Difficulties in executive functioning and impulsivity may contribute to the higher prevalence of disordered eating behaviors in individuals with ADHD (Keller et al., 2024; Martin, Dourish, Hook, Chamberlain, & Higgs, 2022; Nazar et al., 2016), and such behaviors may also serve as coping mechanisms in individuals with neurodevelopmental disorders (e.g., Makin, Zesch, Meyer, Mondelli, & Tchanturia, 2025; Pruccoli et al., 2023). Compared to its prevalence in the general population (1.4%), ADHD is more common among individuals with binge-eating-related disorders, with 26%–31% of these individuals also scoring above the threshold for ADHD (Svedlund et al., 2017). In addition to studies conducted with clinical samples, binge eating behaviours are also associated with self-reported ADHD symptoms (Hanson, Phillips, Hughes, & Corson, 2020). Prospective associations have also been reported between ADHD and binge eating; for example, childhood ADHD (before age 12) predicts binge eating behaviours in adolescence (ages 14–16) and adulthood (over age 18) (Levin & Rawana, 2016; Sonneville et al., 2015).

It is therefore possible that binge eating partially mediates the relationship between ADHD and BMI, with ADHD symptoms potentially leading to higher BMI through binge eating behaviours. Some empirical evidence has provided preliminary support for the mediating role of binge eating in the relationship between ADHD and BMI. For example, in children aged 12 to 15 with obesity, impulsivity was associated with impulsive eating (Nederkoorn, Braet, Van Eijs, Tanghe, & Jansen, 2006). Similarly, a study of adults aged 25 to 46 found that ADHD influences overweight by increasing overeating behaviours, including binge eating (Davis, Levitan, Smith, Tweed, & Curtis, 2006).

It is noteworthy that mechanisms underlying the associations between binge eating and the two core subdomains of ADHD (hyperactivity-impulsivity and inattention) may differ. Both subdomains have been associated with binge eating: inattention with deficits in attentional control and hyperactivity-impulsivity with impaired behavioral control (Tistarelli, Fagnani, Troianiello, Stazi, & Adriani, 2020). However, existing research has primarily focused on the association between hyperactivity-impulsivity and binge eating (e.g., Giel, Teufel, Junne, Zipfel, & Schag, 2017; Schag, Schönleber, Teufel, Zipfel, & Giel, 2013), whereas the role of inattention has received less attention. Inattention subdomain may also be linked to binge eating due to factors such as the impaired perception or utilization of interoceptive signals to guide appetitive behaviour. Supporting this, studies have reported associations between inattention symptoms and disordered eating risk, suggesting that inattention, like hyperactivity–impulsivity, may increase vulnerability to binge eating (E. Martin, Dourish, Rotshtein, Spetter, & Higgs, 2019; Martin, Schell, Srivastav, & Racine, 2020). This highlights the need for further exploration into the distinct roles of different ADHD subdomains in relation to binge eating.

Understanding underlying associations between neurodiversity and disordered eating was a key outcome of lived experience priority-setting exercises (Keller et al., 2024; Thomas, Cooper, & Jones, 2025). The current study aims to explore the predictive role of ADHD in childhood, adolescence, and early adulthood on BMI at age 26, and examine the mediating role of binge eating at age 21. To investigate whether potential mediation effects are confounded by genetic factors, genetically informative mediation models were used (Rosenström et al., 2019). Based on previous research, we hypothesized that binge eating at age 21 will partially mediate the association between ADHD and adult obesity at age 26, after controlling for shared genetic influences.

## Method

This research was pre-registered on the Open Science Framework (OSF) website: OSF | Early ADHD and Obesity: The mediating role of binge eating.

### Participants

The current study used data from the Twins Early Development Study (TEDS), a longitudinal twin cohort following participants from ages 4 to 26 years (Lockhart et al., 2023; Rimfeld et al., 2019; Trouton, Spinath, & Plomin, 2002). TEDS includes twins born in England and Wales between 1994 and 1996. The initial sample comprised 13,926 twin pairs. Participants with severe medical conditions, extreme adverse perinatal circumstances, missing essential background variables, or missing zygosity or gender information were excluded. After screening, 4,363 monozygotic (MZ) and 8,575 dizygotic (DZ) twins remained, with males representing 49.9% of the sample. Over 7,500 twin pairs provided ADHD data in early childhood, and more than 4,500 continued to do so in early adulthood.

### Measures

#### ADHD

Latent factors were specified for ADHD, indicated by multiple measurements (see Table S1). In childhood, ADHD symptoms were measured using the parent-report Strengths and Difficulties Questionnaire (SDQ; Goodman, 1997) at ages 4, 7, and 9, as well as the parent-reported Conners’ Rating Scales-Revised (CRS-R; Conners, Epstein, Angold, & Klaric, 2003) at age 8. During adolescence, ADHD symptoms were assessed using the parent-reported Conners scale and SDQ at ages 12, 14, and 16. During early adulthood (21 years old), ADHD symptoms were assessed with the parent-reported and self-reported SDQ and parent-reported Conners.

We further divided the measured ADHD symptoms into the inattention subdomain and the hyperactivity-impulsivity subdomain based on the content of the questionnaire items. The specific measurements of the ADHD subdomains were consistent with those of overall ADHD. The only exception was the measurement of childhood inattention: to ensure a good fit, it was necessary to exclude the age 4 SDQ. Childhood inattention therefore comprised inattention items from the SDQ at ages 7 and 9, and the Conners at age 8.

### Binge eating

Binge eating symptoms were measured by a brief 12-item questionnaire on eating disorders at 21 years old. This scale was developed by Garner (1991). We selected the 3 items of binge eating subscale in this questionnaire, including “I stuff myself with food”, “I think about binging (overeating)” and “I eat or drink in secrecy”.

### BMI

Height and weight were self-reported at the age of 26, and BMI was calculated using the formula weight (kg)/height (m)² (Bray, 2011).

### Covariates

In this study, we controlled for participants’ sex and age. In addition, given the relative stability of BMI across development, baseline BMI was controlled for in each model: BMI measured at age 4 in the childhood model, age 12 in the adolescent model, and age 21 in the adulthood model.

## Data analysis

### The Phenotypic Mediation Model

Mediation of the association between ADHD and BMI via binge eating was tested in lavaan (version 0.6-20) using models including the direct effect of ADHD on BMI and the indirect effect through binge eating (Figure 1). The indirect effect was estimated as the product of the ADHD → binge eating and binge eating → BMI paths, with significance assessed using 5,000 bootstrap samples. Because the ADHD construct included repeated measures of the same questionnaire (e.g., SDQ) and questionnaires with substantial item overlap (Conners and SDQ), we allowed (1) residual correlations between the Conners and SDQ reported by the same informant at the same time point, and (2) residual correlations between adjacent time points for repeated measurements of the same questionnaire by the same informant. This approach has been supported in previous research (Landis, Edwards, & Cortina, 2009).

**Figure 1.**
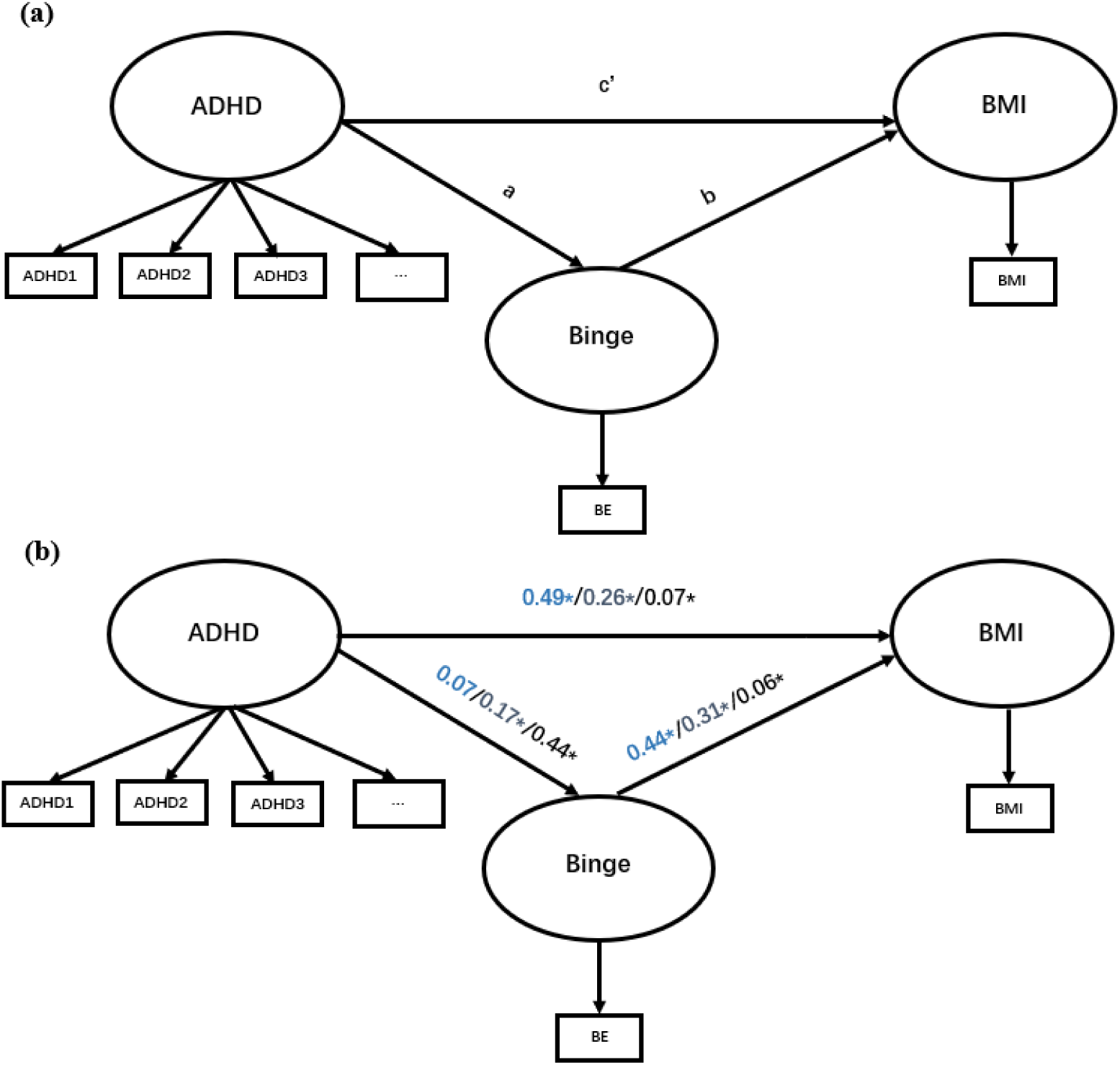
The Phenotypic Factor Mediation Model (a) This model illustrates the mediating role of binge eating in the relationship between ADHD and BMI. ADHD symptoms were assessed using multiple measures at each stage. Binge eating and BMI were measured at ages 21 and 26, respectively. a: direct path from ADHD to binge eating; b: direct path from binge eating to BMI; c’: direct path from ADHD to BMI; Indirect effect = a*b; Total effect = c′ + (a*b). (b) This shows the coefficients for the phenotypic factor mediation models in childhood, adolescence, and adulthood, respectively.

### Multivariate Genetic Model Fitting

In twin studies, monozygotic twins share 100% of their genetic material, whereas dizygotic twins share (on average) 50% of their genetic variance. When phenotypic similarity between monozygotic twins is higher than that of dizygotic twins, it indicates genetic influence (Rijsdijk & Sham, 2002). Therefore, analysis of twin data makes it possible to estimate the extent of genetic and environmental influences on traits.

Specifically, we divided the variances and covariances of ADHD, binge eating, and obesity, as well as the residual variances of their indicator variables, into latent additive genetic components (A), shared environmental components (C), dominant genetic components (D) and non-shared environmental components (E). The A represent the cumulative effects of multiple genetic loci across the genome. The D refers to the effects of alleles that can “mask” or “dominate” the effect of another. The C represents environmental influences that make family members similar to one another, while E includes environmental factors that differentiate family members. MZ twins typically share 100% of their A and dominant genetic components (D), whereas DZ twins typically share 50% of their A and 25% of their D. D components and C components cannot be estimated at the same time in the classical twin design, because when *r*(MZ) > 2\**r*(DZ), the data suggest the influence of D; conversely, when *r*(MZ) < 2\**r*(DZ), it suggests the influence of C (Boomsma, Busjahn, & Peltonen, 2002; Knopik, Neiderhiser, DeFries, & Plomin, 2017).

Previous research in the same sample, using these variables has indicated that ADHD measured at different stages follows an A, D, and E structure (You Oginni, Rijsdijk, Lim, Zavos, & McAdams, 2024). We therefore adopted the A, D, E structure for the ADHD variables in this study. In addition, we also included sibling interaction effects in line with previous research on ADHD (Merwood et al., 2014; Rietveld, Posthuma, & Dolan, 2003; You et al., 2024). For binge eating and BMI, the *r*(MZ) was closer to 2\**r*(DZ), suggesting that the contributions of C or D were small. We further validated the etiological structure of binge eating and BMI within the univariate twin models (see Table S2-S3). The results suggested that in the ACE model, the C components had no significant effects on either binge eating or BMI. Therefore, we tested simplified models in which the C components were removed. After dropping the C components, the simplified models were not significantly different from the full ACE models(p> 0.05). Therefore, in the subsequent analyses, binge eating and BMI were modeled using an A, E structure.

### Biometric Models

To examine the aetiological structure between ADHD, binge eating, and BMI, we constructed several biometric genetic models in OpenMx (version 2.15.5) using full-information maximum likelihood (FIML): a Cholesky decomposition model, an independent pathway model, and a modified biometric mediation model. The first two models specify aetiological overlaps between variables, whereas the latter model includes causal paths. In the Cholesky decomposition model, the first common genetic and environmental factors (A₁ and E_1_) influenced all three traits (ADHD, binge eating, and BMI), indicating their shared genetic and environmental effects. The second common genetic and environmental factors (A₂ and E_2_) influenced binge eating and BMI, representing the residual shared genetic effects between the two traits after accounting for A₁ and E_1_. The third genetic and environmental factors (A₃ and E_3_) influenced only BMI, representing the genetic and environmental effects unique to BMI. In addition, each ADHD measurement was influenced by specific A, D and E components (see Figure 2a). In the independent pathway model, the variables were influenced by a common set of genetic and environmental factors (see Figure 2b). The A and E loaded on all three traits, while each observed variable was also affected by variable-specific A, D_ADHD_, and E components (see Figure 2b). In the modified biometric mediation model, the mediation was built on a three-variable correlation model by adding causal paths between variables. On this basis, a direct path from ADHD to BMI and an indirect path from ADHD to BMI via binge eating were specified (see Figure 2c). Additionally, in the modified biometric mediation model, the correlations in the common A factors among ADHD, binge eating, and BMI were estimated, although shared E was not modeled to allow for the estimation of the mediation paths. Each ADHD measurement was also influenced by specific A, D, and E components (see Figure 2d).

**Figure 2.**
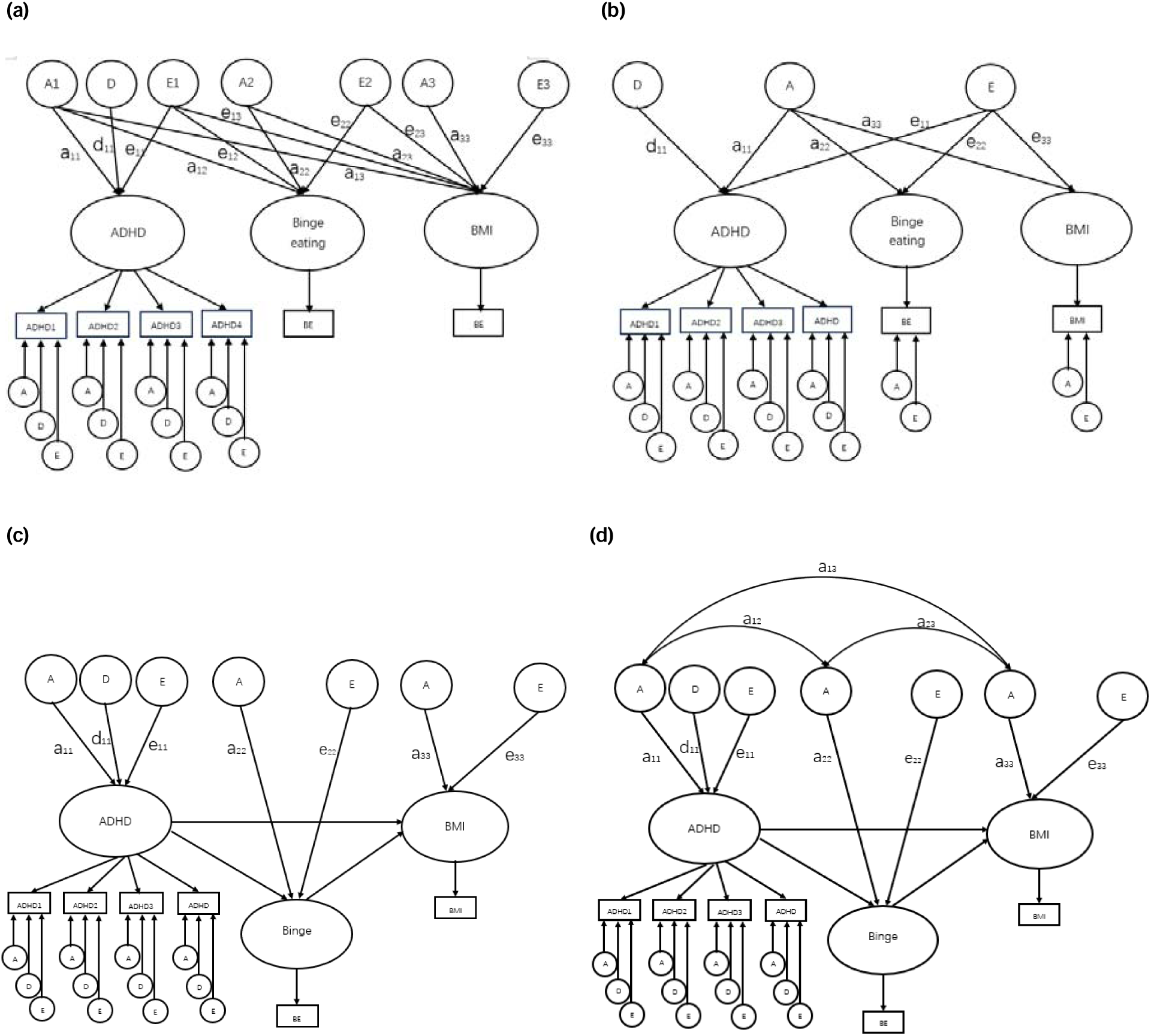
The Biometric Models (a) The Cholesky decomposition model of ADHD, binge eating, and BMI. This model indicates that genetic and environmental effects influence the variables sequentially. (b) The Independent Pathway Model of ADHD, binge eating, and BMI. This model indicates that the variables were influenced by a common set of genetic and environmental factors. (c) The biometric mediation model of ADHD, binge eating, and BMI. This model indicates that ADHD influences BMI through binge eating behaviors. (d) The Modified biometric mediation model of ADHD, binge eating, and BMI. This model indicates that ADHD influences BMI through binge eating behaviors after controlling for shared genetic effects. A: additive genetic components; D: dominant genetic components; E: non-shared environmental components. ADHD symptoms were assessed separately in childhood, adolescence, and adulthood, with each stage incorporating multiple measures. Binge eating was measured at age 21. BMI was measured at age 26.

Model fit was assessed using the Root Mean Square Error of Approximation (RMSEA; Steiger & Lind, 1980), the Tucker-Lewis Index (TLI; Tucker & Lewis, 1973), and the Comparative Fit Index (CFI; Bentler, 1990). A well-fitting model was considered to meet the criteria of RMSEA < 0.08, TLI > 0.90, and CFI > 0.90. Model comparisons for non-nested models were conducted using the Akaike Information Criterion (AIC) and AIC weights (Akaike, 1973, 1974), as well as the-2 Log-Likelihood statistic (-2LL). Generally, a smaller AIC that differs by more than 3 provides some support for a better model fit (Burnham & Anderson, 2004). The comparison of −2LL between the nested models was conducted using a chi-square distribution. A non-significant p-value indicates that, after removing the paths, the more simplified model does not fit the data significantly worse than the original model.

To examine how ADHD in childhood, adolescence, and adulthood predicts BMI through the mediating effect of binge eating at each developmental stage, as well as how genetic and environmental factors influence these mediating effects, above analyses were repeated separately for each developmental period. Additionally, we conducted the exploratory analyses separately for the two ADHD subdomains, inattention and hyperactivity-impulsivity.

## Results

### Descriptive statistics

Descriptive statistics of the observed variables are shown in Table S4. In general, symptoms of parent reported ADHD decreased with age from mean of 4.02 to 1.96 from ages 4 to 21 respectively. The average binge eating behavior score, measured at age 21, was 3.75. The mean BMI in adulthood (age 26) was 24.74, with 23.0% of participants classified as overweight (25 ≤ BMI < 30) and 13.8% classified as obese (BMI ≥ 30). Phenotypic correlations among ADHD, binge eating and BMI are shown in Table S5. ADHD symptoms in adulthood (age 21) showed the highest correlation with binge eating at age 21 (*r* = 0.25), while childhood ADHD symptoms (*r* = 0.06) and adolescent ADHD symptoms (*r* = 0.10) had significantly lower correlations with binge eating at age 21. Therefore, overall correlations between ADHD symptoms and binge eating across all time points were small, with only a slight increase between symptoms measured at the same time point. ADHD symptoms at each age also showed significant but small correlations with BMI at age 26 (*r* = 0.11 to 0.18). The correlation between binge eating and BMI was 0.24.

### Phenotypic mediation models

In the results of the phenotypic mediation models (see Tables 1), the paths from ADHD to binge eating were not significant in childhood, but were significant in adolescence (estimate = 0.17, 95% CI 0.10 to 0.25) and adulthood (estimate = 0.44, 95% CI 0.38 to 0.53). Paths from ADHD to BMI were significant at all three stages, with the estimates of 0.49 (95% CI 0.30 to 0.70) in childhood, 0.26 (95% CI 0.15 to 0.38) in adolescence, and 0.07 (95% CI 0.02 to 0.13) in adulthood. The indirect associations between ADHD and adult BMI through binge eating were significant in adolescence and adulthood, but not childhood. Specifically, during adolescence, the mediated effect was 0.05 (95% CI 0.03 to 0.08); during adulthood, the mediated effect was 0.03 (95% CI 0.01 to 0.05).

### Biometric models

Table S5 presents the estimates of A, D, and E for each variable from the best-fitting models, and Table S3 shows the estimates of A and E for binge eating and BMI from the univariate twin models. Specifically, ADHD symptoms were consistently influenced by genetic factors. In childhood, dominant genetic effects were the primary influence (d² = 0.75), whereas in adolescence and adulthood, additive genetic influences increased (h² = 0.73 and 0.43, respectively). The proportion of variance accounted for by non-shared environmental factors was small to moderate from childhood to adulthood (e² = 0.14–0.26). In addition, binge eating was influenced by substantial environmental effects (e² ≈ 0.67), along with moderate additive genetic effects (h² ≈ 0.33). For BMI, in the estimates from the univariate twin models, additive genetic effects were the primary influence (h² = 0.75), while the proportion of variance accounted for by non-shared environmental factors was small (e² = 0.25). The heritability estimates for BMI from the best-fitting multivariate models (Table S5) were somewhat reduced compared to univariate estimates as BMI at the previous time-point was controlled for.

The results of model comparisons are shown in Table 1, Table 3 and Table S6. The AIC differences between the modified biometric mediation model (childhood: AIC = 264124; adolescence: AIC = 267719) and the Cholesky decomposition model (childhood: AIC = 264124; adolescence: AIC = 267720) were less than 1 in childhood and adolescence. This suggests no strong evidence to support either model. In the modified biometric mediation models, genetic correlations were observed among the variables. After accounting for genetic overlap, only the paths from binge eating to BMI were significant. The Cholesky decomposition model makes no assumptions about causal relationships between variables; based on parsimony, it was selected as the best-fitting model. This model included significant shared genetic components among ADHD, binge eating, and BMI. Specifically, the first genetic factor (A_₁_) significantly loaded on ADHD (childhood: a_11_ = 0.03, 95% CI 0.02 to 0.05; adolescence: a_11_ = 0.73, 95% CI 0.62 to 0.84), binge eating (childhood: a_12_ = 0.16, 95% CI 0.06 to 0.26; adolescence: a_12_ = 0.01, 95% CI 0.01 to 0.02), and BMI (childhood: a_13_ = 0.63, 95% CI 0.42 to 0.74; adolescence: a_13_ = 0.04, 95% CI 0.02 to 0.06). The second genetic factor (A_₂_) influenced binge eating (childhood: a_22_ = 0.17, 95% CI 0.07 to 0.28; adolescence: a_22_ = 0.32, 95% CI 0.27 to 0.36) and BMI (childhood: a_23_ = 0.11, 95% CI 0.004 to 0.31; adolescence: a_23_ = 0.05, 95% CI 0.02 to 0.09), representing the residual shared genetic effects between the two after accounting for A_₁_. Apart from A_₁_ and A_₂_, no unique genetic effects (A_₃_) on BMI were observed in childhood, whereas it was still observed in adolescence (a_33_ = 0.54, 95% CI 0.49 to 0.58). Regarding environmental influences, common non-shared environmental influences among the traits were small and non-significant. This indicates that the E on the variables were independent of each other, with estimates of 21% (95% CI 0.20 to 0.23), 67% (95% CI 0.63 to 0.71), and 26% (95% CI 0.23 to 0.29) for ADHD, binge eating, and BMI in childhood, respectively, and 14% (95% CI 0.13 to 0.15), 67% (95% CI 0.63 to 0.71), and 36% (95% CI 0.33 to 0.40) in adolescence. Therefore, the associations between childhood and adolescent ADHD, adulthood binge eating, and BMI were largely explained by common genetic factors, although the pathway from binge eating to BMI may also be significant.

**Table 1.**
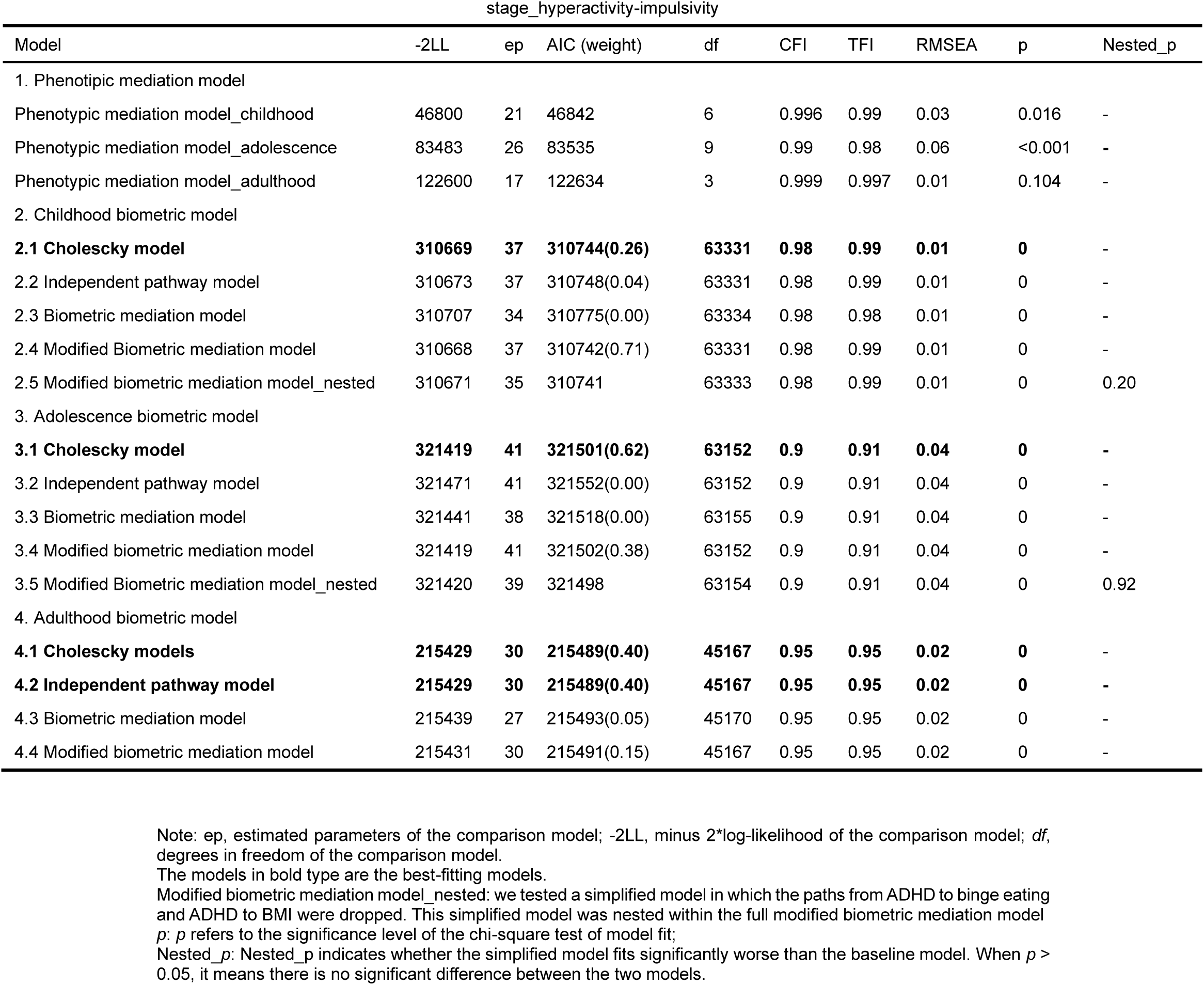
Fit statistics for Cholesky decomposition models, Independent Pathway Models and Adjusted biometric mediation models at each stage_hyperactivity-impulsivity.

| stage_hyperactivity-impulsivity |  |  |  |  |  |  |  |  |  |
| --- | --- | --- | --- | --- | --- | --- | --- | --- | --- |
| Model | -2LL | ep | AIC (weight) | df | CFI | TFI | RMSEA | p | Nested_p |
| 1. Phenotypic mediation model |  |  |  |  |  |  |  |  |  |
| Phenotypic mediation model_childhood | 46800 | 21 | 46842 | 6 | 0.996 | 0.99 | 0.03 | 0.016 | - |
| Phenotypic mediation model_adolescence | 83483 | 26 | 83535 | 9 | 0.99 | 0.98 | 0.06 | <0.001 | - |
| Phenotypic mediation model_adulthood | 122600 | 17 | 122634 | 3 | 0.999 | 0.997 | 0.01 | 0.104 | - |
| 2. Childhood biometric model |  |  |  |  |  |  |  |  |  |
| **2.1 Cholesky model** | **310669** | **37** | **310744(0.26)** | **63331** | **0.98** | **0.99** | **0.01** | **0** | - |
| 2.2 Independent pathway model | 310673 | 37 | 310748(0.04) | 63331 | 0.98 | 0.99 | 0.01 | 0 | - |
| 2.3 Biometric mediation model | 310707 | 34 | 310775(0.00) | 63334 | 0.98 | 0.98 | 0.01 | 0 | - |
| 2.4 Modified Biometric mediation model | 310668 | 37 | 310742(0.71) | 63331 | 0.98 | 0.99 | 0.01 | 0 | - |
| 2.5 Modified biometric mediation model_nested | 310671 | 35 | 310741 | 63333 | 0.98 | 0.99 | 0.01 | 0 | 0.20 |
| 3. Adolescence biometric model |  |  |  |  |  |  |  |  |  |
| **3.1 Cholesky model** | **321419** | **41** | **321501(0.62)** | **63152** | **0.9** | **0.91** | **0.04** | **0** | - |
| 3.2 Independent pathway model | 321471 | 41 | 321552(0.00) | 63152 | 0.9 | 0.91 | 0.04 | 0 | - |
| 3.3 Biometric mediation model | 321441 | 38 | 321518(0.00) | 63155 | 0.9 | 0.91 | 0.04 | 0 | - |
| 3.4 Modified biometric mediation model | 321419 | 41 | 321502(0.38) | 63152 | 0.9 | 0.91 | 0.04 | 0 | - |
| 3.5 Modified Biometric mediation model_nested | 321420 | 39 | 321498 | 63154 | 0.9 | 0.91 | 0.04 | 0 | 0.92 |
| 4. Adulthood biometric model |  |  |  |  |  |  |  |  |  |
| **4.1 Cholesky models** | **215429** | **30** | **215489(0.40)** | **45167** | **0.95** | **0.95** | **0.02** | **0** | - |
| **4.2 Independent pathway model** | **215429** | **30** | **215489(0.40)** | **45167** | **0.95** | **0.95** | **0.02** | **0** | - |
| 4.3 Biometric mediation model | 215439 | 27 | 215493(0.05) | 45170 | 0.95 | 0.95 | 0.02 | 0 | - |
| 4.4 Modified biometric mediation model | 215431 | 30 | 215491(0.15) | 45167 | 0.95 | 0.95 | 0.02 | 0 | - |

Note: ep, estimated parameters of the comparison model; -2LL, minus 2\*log-likelihood of the comparison model; *df*, degrees in freedom of the comparison model.
The models in bold type are the best-fitting models.
Modified biometric mediation model\_nested: we tested a simplified model in which the paths from ADHD to binge eating and ADHD to BMI were dropped. This simplified model was nested within the full modified biometric mediation model
*p*: *p* refers to the significance level of the chi-square test of model fit;
Nested\_p: Nested\_p indicates whether the simplified model fits significantly worse than the baseline model. When *p* > 0.05, it means there is no significant difference between the two models.

**Table 2.**
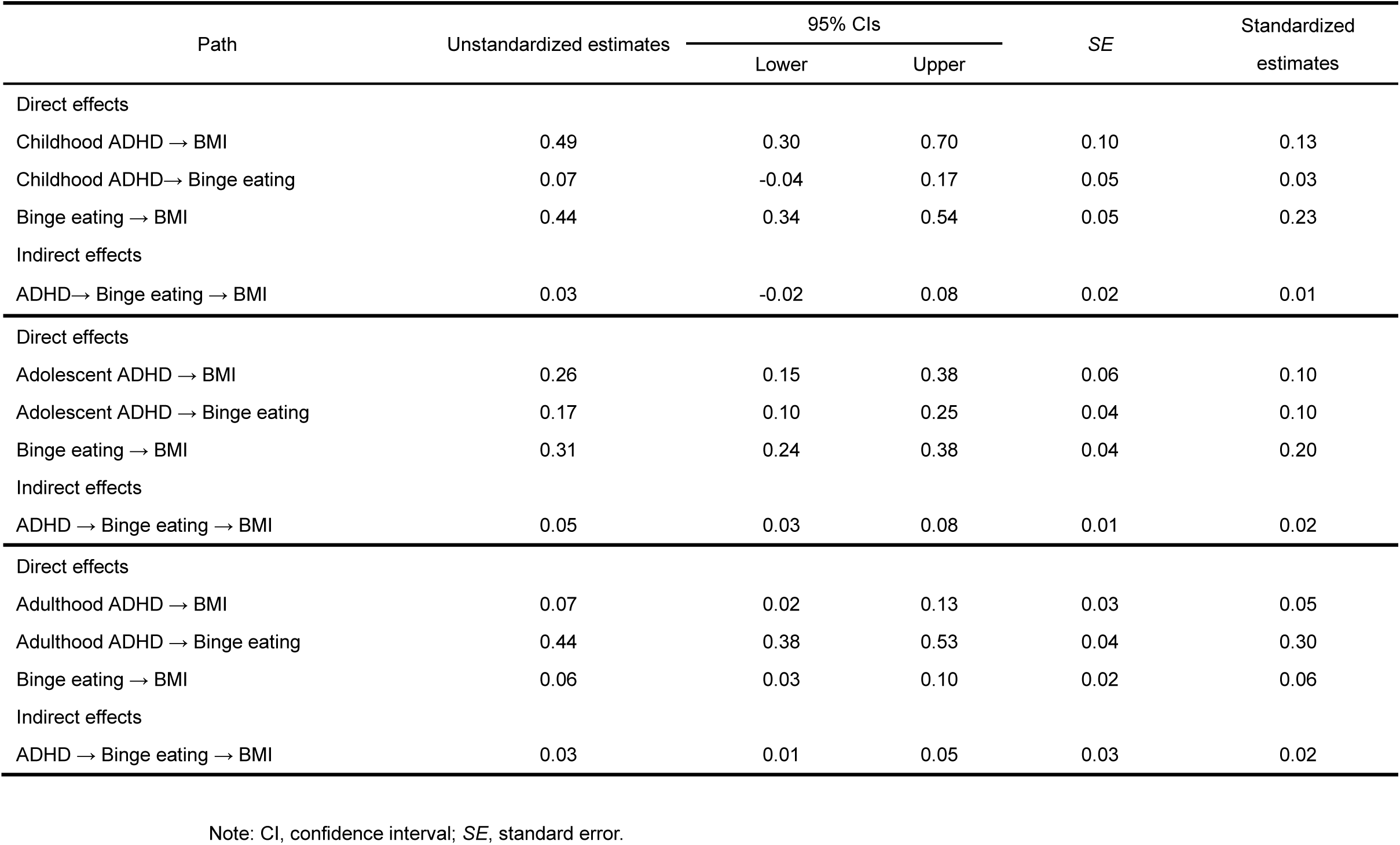
The phenotypic mediation model of ADHD, binge eating, and BMI.

| Path | Unstandardized estimates | 95% CIs |  | SE | Standardized estimates |
| --- | --- | --- | --- | --- | --- |
|  |  | Lower | Upper |  |  |
| Direct effects |  |  |  |  |  |
| Childhood ADHD → BMI | 0.49 | 0.30 | 0.70 | 0.10 | 0.13 |
| Childhood ADHD→ Binge eating | 0.07 | -0.04 | 0.17 | 0.05 | 0.03 |
| Binge eating → BMI | 0.44 | 0.34 | 0.54 | 0.05 | 0.23 |
| Indirect effects |  |  |  |  |  |
| ADHD→ Binge eating → BMI | 0.03 | -0.02 | 0.08 | 0.02 | 0.01 |
| Direct effects |  |  |  |  |  |
| Adolescent ADHD → BMI | 0.26 | 0.15 | 0.38 | 0.06 | 0.10 |
| Adolescent ADHD → Binge eating | 0.17 | 0.10 | 0.25 | 0.04 | 0.10 |
| Binge eating → BMI | 0.31 | 0.24 | 0.38 | 0.04 | 0.20 |
| Indirect effects |  |  |  |  |  |
| ADHD → Binge eating → BMI | 0.05 | 0.03 | 0.08 | 0.01 | 0.02 |
| Direct effects |  |  |  |  |  |
| Adulthood ADHD → BMI | 0.07 | 0.02 | 0.13 | 0.03 | 0.05 |
| Adulthood ADHD → Binge eating | 0.44 | 0.38 | 0.53 | 0.04 | 0.30 |
| Binge eating → BMI | 0.06 | 0.03 | 0.10 | 0.02 | 0.06 |
| Indirect effects |  |  |  |  |  |
| ADHD → Binge eating → BMI | 0.03 | 0.01 | 0.05 | 0.03 | 0.02 |

Note: CI, confidence interval; SE, standard error.

**Table 3.**
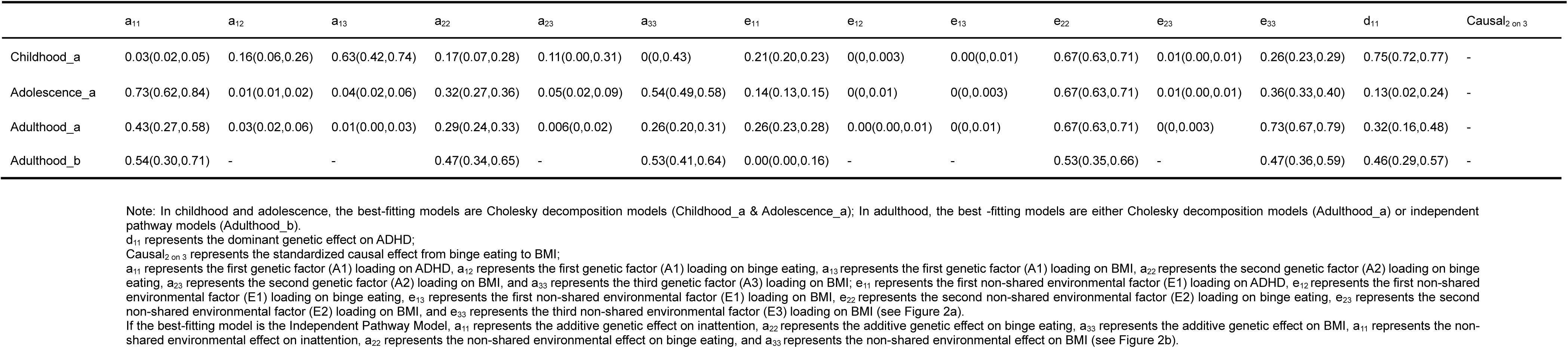
Parameter estimates (95% confidence intervals) in every stage from the best-fitting model.

In adulthood, the Cholesky decomposition model and the independent pathway model with AICs of 215429 demonstrated the best fit to the data (see Tables 1 and 3). Specifically, in the Cholesky decomposition model, a common genetic factor (A_₁_) significantly influenced ADHD (a_11_ = 0.43, 95% CI 0.27 to 0.58), binge eating (a_12_ = 0.03, 95% CI 0.02 to 0.06), and BMI (a_13_ = 0.01, 95% CI 0.001 to 0.03). In addition, another common genetic factor (A_₂_) was identified, representing the residual shared genetic effects between binge eating (a_22_ = 0.29, 95% CI 0.24 to 0.33) and BMI (a_23_ = 0.01, 95% CI 0.00 to 0.02) after accounting for A_₁_. Finally, there was also a significant genetic effect (A_₃_) that influenced only BMI (a_33_ = 0.26, 95% CI 0.20 to 0.31). Regarding environmental influences, it accounted for 26% (95% CI 0.23 to 0.28), 67% (95% CI 0.63 to 0.71), and 73% (95% CI 0.67 to 0.79) of the variance in ADHD, binge eating, and BMI, respectively. While the common non-shared environmental influences among the traits were small and non-significant. In the independent pathway model, common genetic and environmental factors influenced all three traits. The common genetic factor (A) explained 21% (95% CI 0.15–0.27), 32% (95% CI 0.23–0.41), and 25% (95% CI 0.20–0.31) of the variance in inattention, binge eating, and BMI, respectively. The common non-shared environmental factor (C) accounted for 14% (95% CI 0.13–0.15), 67% (95% CI 0.63–0.71), and 37% (95% CI 0.33–0.41) of the variance in ADHD, binge eating, and BMI. Overall, the best adulthood models, after accounting for shared genetic and environmental influences, included no paths between the phenotypes.

### Models of ADHD subdomains

Phenotypic mediation models among ADHD subdomains, binge eating, and BMI showed significant mediating effects in line with results for the overall ADHD measure. However, in the biometric models, no mediating effects were found. The associations between ADHD subdomains and binge eating, as well as between ADHD subdomains and BMI, were primarily explained by common genetic factors. For details, please refer to the supplementary materials (see Supplementary Text S1-S2; Table S7-Table S16).

## Discussion

This study is the first to explore how ADHD symptoms at different developmental stages are associated with adult BMI, with a particular focus on the mediating role of binge eating. At the phenotypic level, our mediation model indicated that ADHD in adolescence and adulthood influenced subsequent BMI via binge eating, but this mediating effect was no longer significant after accounting for shared genetic factors. These results suggest that the observed phenotypic mediation effects might be largely due to shared genetic influences underlying these three traits.

In the phenotypic mediation models, significant direct effects from ADHD, hyperactivity–impulsivity, and inattention to binge eating were observed in adolescence and adulthood. This may be because, during childhood, diet is more strongly influenced by parents, whereas from adolescence onward, food choice transitions from primarily depending on caregivers to adolescents taking more responsibility for food acquisition and preparation (Neufeld et al., 2022). Therefore, fewer instances of binge eating associated with ADHD symptoms are observed during childhood, with such associations becoming more apparent only after adolescence begins. In addition, except for inattention in adulthood, the direct effects of ADHD and ADHD subdomains on BMI were significant across developmental stages. These findings are consistent with previous studies suggesting that individuals with ADHD may experience more difficulties with impulse inhibition, which has been linked to disordered eating behaviors and obesity (e.g., Davis et al., 2006; Nederkoorn et al., 2006). Moreover, our results indicate that inattention, and not just hyperactivity–impulsivity, predicts binge eating, potentially reflecting deficits in attentional control could also impair the regulation of eating behavior. Individuals with ADHD are more likely to be influenced by immediate reinforcement contingencies in their environment (Tripp & Wickens, 2008), which is associated with the difficulty in sustaining attention. Binge eating can provide immediate reward, which may explain why individuals with higher levels of inattention symptoms are more likely to engage in binge eating as a form of immediate reinforcement. Furthermore, we observed that binge eating mediated the pathway from adolescence and early adulthood ADHD to BMI. This finding may be interpreted in a traditional mediation model as indicating that earlier ADHD symptoms continue to influence BMI at age 26 through binge eating behaviors in early adulthood. This highlights the longitudinal associations between ADHD symptoms, eating behaviors, and BMI across developmental stages.

In the biometric models, shared genetic risk (e.g., Capusan et al., 2017; Cortese & Peñalver, 2010; Karhunen et al., 2021) contributed to correlations between early ADHD symptoms and subsequent binge eating and BMI. This suggests that the mediation effects observed in the phenotypical models may in fact reflect confounding by shared genetic factors. These results differ from previous research using mendelian randomization (Chen et al., 2023; Liu et al., 2021) which reported a bidirectional causal association between ADHD and BMI. Several factors may explain the discrepancies between our results and these two studies. First, methodological differences may be a reason for the discrepancy between the results from twin models and several mendelian randomization studies. Mendelian randomization uses genetic liability to ADHD as an instrumental variable to test potential causal effects of ADHD on BMI, whereas the modified biometric mediation models in the present study focus on associations within a specific developmental period (ages 4 to 26). Therefore, the two approaches may not yield consistent results due to differences in measurement and modeling. In addition, our twin models with directional paths examine prospective phenotypic associations (e.g., ADHD at ages 7, 12, or 21 predicting BMI at 26). If the effect of ADHD on BMI is largely immediate rather than a long-term accumulation across developmental stages, these twin models may fail to detect a significant pathway. This might further imply that, if early ADHD symptoms remit, their long-term impact on later psychiatric symptoms and developmental outcomes is relatively small. These results are echoed in previous research, which suggests that, compared with individuals with persistent ADHD, those whose symptoms remit experience fewer psychiatric symptoms later in life (e.g., Agnew-Blais et al., 2016; Agnew-Blais et al., 2018), highlighting that remitting symptoms are associated with a milder course and fewer long-term impacts.

The current study makes several important contributions. A key strength is the developmental perspective taken. We used the same sample from childhood to adulthood to examine how ADHD symptoms at different stages influence subsequent eating behaviors and BMI. Secondly, this study demonstrates that the mediating pathway, frequently reported in previous research, whereby early ADHD influences BMI through impaired impulse control over food, may in fact be confounded by genetic factors rather than representing a true symptom-level mediation. This provides new insights into the nature of the comorbidity between ADHD, binge eating and BMI.

Nevertheless, this study also has some limitations. For example, in many longitudinal studies, including the TEDS database, individuals with more severe early symptoms were more likely to drop out over time (Rimfeld et al., 2019; Teague et al., 2018), and this attrition may also have contributed to the absence of detectable mediation effects in the modified biometric mediation models. Moreover, the measure of eating behaviors used in the current study consisted of three items assessing binge eating. However, there may be other eating behaviors that link ADHD and BMI. A person with poor impulse control may be more prone to eat more than they need even if they do not binge eat, and such overeating without loss of control has also been associated with weight status (Neumark-Sztainer et al., 2015). ADHD may make the effort to resist food more difficult, thereby increasing the likelihood of becoming overweight without the need for binge eating as a mediator. Therefore, future research could further examine how ADHD influences other types of eating behaviors apart from binge eating.

## Conclusion

This study investigated the associations between ADHD symptoms at different developmental stages, binge eating at age 21, and BMI at age 26. The results showed that in the phenotypic mediation model, adolescent and adulthood ADHD influenced subsequent BMI through the mediation of binge eating. However, this mediation effect was in fact attributable to confounding by shared genetic factors in biometric models. Thus, the associations between ADHD and binge eating, as well as between ADHD and BMI, are explained by shared genetic effects. This twin study reveals shared liability across ADHD, binge eating, and BMI. Intervention strategies could focus more on common underlying behavioural and self-regulatory mechanisms across these traits, as well as placing more emphasis on symptom patterns within families.

## Data Availability

I used datasets or databases (TEDS) that were obtained after a request, or required application.

## Acknowledgements

The authors gratefully acknowledge the ongoing contribution of the TEDS participants and their families.

## Financial support

YY is supported by China Scholarship Council; TAM is supported by a Wellcome Trust Senior Research Fellowship awarded to TAM (Grant 220382/Z/20/Z). TEDS is supported by a programme grant (MR/V012878/1) to Professor Thalia Eley from the UK Medical Research Council (previously MR/M021475/1 awarded to Professor Robert Plomin). The authors gratefully acknowledge the ongoing contribution of the TEDS participants and their families.

## Ethical standards

The authors assert that all procedures contributing to this work comply with the ethical standards of the relevant national and institutional committees on human experimentation and with the Helsinki Declaration of 1975, as revised in 2008.

## Competing Interests

The authors declare none.

## Notes

### Competing Interest Statement

The authors have declared no competing interest.

### Author Declarations

I used datasets or databases (TEDS) that were obtained after a request, or required application. All datasets used in this study were de-identified before being accessed by the researchers.

## References

Agnew-Blais, J. C., Polanczyk, G., Danese, A., Wertz, J., Moffitt, T. E., & Arseneault, L. (2016). Persistence, remission and emergence of ADHD in young adulthood: results from a longitudinal, prospective population-based cohort. JAMA psychiatry, 73(7), 713–720. 10.1001/jamapsychiatry.2016.0465

Agnew-Blais, J. C., Polanczyk, G. V., Danese, A., Wertz, J., Moffitt, T. E., & Arseneault, L. (2018). Young adult mental health and functional outcomes among individuals with remitted, persistent and late-onset ADHD. The British journal of psychiatry, 213(3), 526–534. 10.1192/bjp.2018.97

Akaike, H. (1974). A new look at the statistical model identification. IEEE transactions on automatic control, 19(6), 716–723. 10.1109/TAC.1974.1100705

Akaike, H. (1973). Information theory and an extension of the maximum likelihood principle. In B. N. Pet rov & F. Caski (Eds.), Proceedings of the Second International Symposium on Information Theory (pp. 267–281). Budapest: Akademiai Kiado

American Psychiatric Association. (2013). Diagnostic and statistical manual of mental disorders (5th ed.). Arlington, VA: American Psychiatric Publishing.

Bentler, P. M. (1990). Comparative fit indexes in structural models. Psychological Bulletin, 107(2), 238–246. 10.1037/0033-2909.107.2.238

Boomsma, D., Busjahn, A., & Peltonen, L. (2002). Classical twin studies and beyond. Nature reviews genetics, 3(11), 872–882. 10.1038/nrg932

Bray, G. A. (2011). A guide to obesity and the metabolic syndrome. Boca Raton, Florida: CRC Press.

Burnham, K. P., & Anderson, D. R. (2004). Multimodel inference: Understanding AIC and BIC in model selection. Sociological Methods & Research, 33(2), 261–304. 10.1177/0049124104268644

Capusan, A. J., Yao, S., Kuja-Halkola, R., Bulik, C. M., Thornton, L. M., Bendtsen, P., & Larsson, H. (2017). Genetic and environmental aspects in the association between attention-deficit hyperactivity disorder symptoms and binge-eating behavior in adults: a twin study. Psychological medicine, 47(16), 2866–2878. 10.1017/S0033291717001416

Chen, W., Feng, J., Jiang, S., Guo, J., Zhang, X., Zhang, X.,…& Dong, Z. (2023). Mendelian randomization analyses identify bidirectional causal relationships of obesity with psychiatric disorders. Journal of Affective Disorders, 339, 807–814. 10.1016/j.jad.2023.07.044

Conners, C. K., Epstein, J. N., Angold, A., & Klaric, J. (2003). Continuous performance test performance in a normative epidemiological sample. Journal of abnormal child psychology, 31, 555–562. 10.1023/A:1025457300409

Cortese, S., Bernardina, B. D., & Mouren, M. C. (2007). Attention-deficit/hyperactivity disorder (ADHD) and binge eating. Nutrition reviews, 65(9), 404–411. 10.1111/j.1753-4887.2007.tb00318.x

Cortese, S., & Peñalver, C. M. (2010). Comorbidity between ADHD and obesity: exploring shared mechanisms and clinical implications. Postgraduate medicine, 122(5), 88–96. 10.3810/pgm.2010.09.2205

Cortese, S., Moreira-Maia, C. R., St. Fleur, D., Morcillo-Peñalver, C., Rohde, L. A., & Faraone, S. V. (2016). Association between ADHD and obesity: a systematic review and meta-analysis. American journal of psychiatry, 173(1), 34–43. 10.1176/appi.ajp.2015.1502026

Davis, C., Levitan, R. D., Smith, M., Tweed, S., & Curtis, C. (2006). Associations among overeating, overweight, and attention deficit/hyperactivity disorder: a structural equation modelling approach. Eating behaviors, 7(3), 266–274. 10.1016/j.eatbeh.2005.09.006

de Zwaan, M. (2001). Binge eating disorder and obesity. International journal of obesity, 25(1), S51–S55. 10.1038/sj.ijo.0801699

Fliers, E. A., Buitelaar, J. K., Maras, A., Bul, K., Höhle, E., Faraone, S. V., & Rommelse, N. N. (2013). ADHD is a risk factor for overweight and obesity in children. Journal of Developmental & Behavioral Pediatrics, 34(8), 566–574. 10.1097/DBP.0b013e3182a50a67

Fuemmeler, B. F., Østbye, T., Yang, C., McClernon, F. J., & Kollins, S. H. (2011). Association between attention-deficit/hyperactivity disorder symptoms and obesity and hypertension in early adulthood: a population-based study. International journal of obesity, 35(6), 852–862. 10.1038/ijo.2010.214

Garner, D. M. (1991). Eating disorder inventory–2 (EDI-2): Professional manual. Odessa, FL: Psychological Assessment Resources.

Giel, K. E., Teufel, M., Junne, F., Zipfel, S., & Schag, K. (2017). Food-related impulsivity in obesity and binge eating disorder—a systematic update of the evidence. Nutrients, 9(11), 1170. 10.3390/nu9111170

Goodman, R. (1997). The Strengths and Difficulties Questionnaire: a research note. Journal of child psychology and psychiatry, 38(5), 581–586. 10.1111/j.1469-7610.1997.tb01545.x

Hanson, J. A., Phillips, L. N., Hughes, S. M., & Corson, K. (2020). Attention-deficit hyperactivity disorder symptomatology, binge eating disorder symptomatology, and body mass index among college students. Journal of American College Health, 68(5), 543–549. 10.1080/07448481.2019.1583651

Kaisari, P., Dourish, C. T., & Higgs, S. (2017). Attention deficit hyperactivity disorder (ADHD) and disordered eating behaviour: a systematic review and a framework for future research. Clinical psychology review, 53, 109–121. 10.1016/j.cpr.2017.03.002

Karhunen, V., Bond, T. A., Zuber, V., Hurtig, T., Moilanen, I., Järvelin, M. R.,…& Rodriguez, A. (2021). The link between attention deficit hyperactivity disorder (ADHD) symptoms and obesity-related traits: genetic and prenatal explanations. Translational Psychiatry, 11(1), 455. 10.1038/s41398-021-01584-4

Keller, J., Herle, M., Mandy, W., & Leno, V. C. (2024). The overlap of disordered eating, autism and ADHD: future research priorities as identified by adults with lived experience. The Lancet Psychiatry, 11(12), 1030–1036. 10.1016/S2215-0366(24)00186-X

Knopik, V. S., Neiderhiser, J. M., DeFries, J. C., & Plomin, R. (2017). Behavioral genetics. New York, NY: Worth Publishers, Macmillan Learning.

Landis, R. S., Edwards, B. D., & Cortina, J. M. (2009). Correlated residuals among items in the estimation of measurement models. In C. E. Lance & R. J. Vandenberg (Eds.), Statistical and methodological myths and urban legends: Doctrine, verity, and fable in the organizational and social sciences (pp. 195–214). New York, NY: Routledge.

Liu, C. Y., Schoeler, T., Davies, N. M., Peyre, H., Lim, K. X., Barker, E. D.,…& Pingault, J. B. (2021). Are there causal relationships between attention-deficit/hyperactivity disorder and body mass index? Evidence from multiple genetically informed designs. International Journal of Epidemiology, 50(2), 496–509. 10.1093/ije/dyaa214

Levin, R. L., & Rawana, J. S. (2016). Attention-deficit/hyperactivity disorder and eating disorders across the lifespan: a systematic review of the literature. Clinical psychology review, 50, 22–36. 10.1016/j.cpr.2016.09.010

Lockhart, C., Bright, J., Ahmadzadeh, Y., Breen, G., Bristow, S., Boyd, A., & Eley, T. C. (2023). Twins Early Development Study (TEDS): A genetically sensitive investigation of mental health outcomes in the mid_-_twenties. JCPP advances, 3(2), e12154. 10.1002/jcv2.12154

Makin, L., Zesch, E., Meyer, A., Mondelli, V., & Tchanturia, K. (2025). Autism, ADHD, and Their Traits in Adults With Bulimia Nervosa and Binge Eating Disorder: A Scoping Review. European Eating Disorders Review. 1–19. 10.1002/erv.3177

Martin, E., Dourish, C. T., Hook, R., Chamberlain, S. R., & Higgs, S. (2022). Associations between inattention and impulsivity ADHD symptoms and disordered eating risk in a community sample of young adults. Psychological medicine, 52(13), 2622–2631. https://doi/org/10.1017/S0033291720004638

Martin, S. J., Schell, S. E., Srivastav, A., & Racine, S. E. (2020). Dimensions of unhealthy exercise and their associations with restrictive eating and binge eating. Eating behaviors, 39, 101436. 10.1016/j.eatbeh.2020.101436

Martin, E., Dourish, C. T., Rotshtein, P., Spetter, M. S., & Higgs, S. (2019). Interoception and disordered eating: A systematic review. Neuroscience & Biobehavioral Reviews, 107, 166–191. 10.1016/j.neubiorev.2019.08.020

Merwood, A., Chen, W., Rijsdijk, F., Skirrow, C., Larsson, H., Thapar, A., & Asherson, P. (2014). Genetic associations between the symptoms of attention-deficit/hyperactivity disorder and emotional lability in child and adolescent twins. Journal of the American Academy of Child & Adolescent Psychiatry, 53(2), 209–220. 10.1016/j.jaac.2013.11.006

Nazar, B. P., de Sousa Pinna, C. M., Suwwan, R., Duchesne, M., Freitas, S. R., Sergeant, J., & Mattos, P. (2016). ADHD rate in obese women with binge eating and bulimic behaviors from a weight-loss clinic. Journal of Attention Disorders, 20(7), 610–616. 10.1177/1087054712455503

Nederkoorn, C., Braet, C., Van Eijs, Y., Tanghe, A., & Jansen, A. (2006). Why obese children cannot resist food: the role of impulsivity. Eating behaviors, 7(4), 315–322. 10.1016/j.eatbeh.2005.11.005

Neumark-Sztainer, D., Berge, J., Pisetsky, E., MacLehose, R., Loth, K., & Goldschmidt, A. (2015). Overeating with and Without Loss of Control: Associations with Weight Status, Weight-Related Characteristics, and Psychosocial Health. International Journal of Eating Disorders, 48(8), 1150–1157. 10.1002/eat.22465

Neufeld, L. M., Andrade, E. B., Suleiman, A. B., Barker, M., Beal, T., Blum, L. S.,…& Zou, Z. (2022). Food choice in transition: adolescent autonomy, agency, and the food environment. The lancet, 399(10320), 185–197. 10.1016/S0140-6736(21)01687-1

Nigg, J. T., Johnstone, J. M., Musser, E. D., Long, H. G., Willoughby, M. T., & Shannon, J. (2016). Attention-deficit/hyperactivity disorder (ADHD) and being overweight/obesity: New data and meta-analysis. Clinical psychology review, 43, 67–79. 10.1016/j.cpr.2015.11.005

Pingault, J. B., Viding, E., Galéra, C., Greven, C. U., Zheng, Y., Plomin, R., & Rijsdijk, F. (2015). Genetic and environmental influences on the developmental course of attention-deficit/hyperactivity disorder symptoms from childhood to adolescence. JAMA psychiatry, 72(7), 651–658. 10.1001/jamapsychiatry.2015.0469

Pruccoli, J., Guardi, G., La Tempa, A., Valeriani, B., Chiavarino, F., & Parmeggiani, A. (2023). Food and development: Children and adolescents with neurodevelopmental and comorbid eating disorders—A case series. Behavioral Sciences, 13(6), 499. 10.3390/bs13060499

Rietveld, M. J. H., Posthuma, D., & Dolan, C. V. (2003). ADHD: Sibling interaction or dominance: An evaluation of statistical power. Behavior Genetics, 33(3), 247–255. 10.1023/A:1023490307170

Rijsdijk, F. V., & Sham, P. C. (2002). Analytic approaches to twin data using structural equation models. Briefings in bioinformatics, 3(2), 119–133. 10.1093/bib/3.2.119

Rimfeld, K., Malanchini, M., Spargo, T., Spickernell, G., Selzam, S., McMillan, A.,…& Plomin, R. (2019). Twins early development study: A genetically sensitive investigation into behavioral and cognitive development from infancy to emerging adulthood. Twin Research and Human Genetics, 22(6), 508–513. 10.1017/thg.2019.56

Rosenström, T., Czajkowski, N. O., Ystrom, E., Krueger, R. F., Aggen, S. H., Gillespie, N. A., & Torvik, F. A. (2019). Genetically informative mediation modeling applied to stressors and personality-disorder traits in etiology of alcohol use disorder. Behavior Genetics, 49, 11–23. 10.1007/s10519-018-9941-z

Schag, K., Schönleber, J., Teufel, M., Zipfel, S., & Giel, K. E. (2013). Food_-_related impulsivity in obesity and Binge Eating Disorder–a systematic review. Obesity reviews, 14(6), 477–495. 10.1111/obr.12017

Sonneville, K. R., Calzo, J. P., Horton, N. J., Field, A. E., Crosby, R. D., Solmi, F., & Micali, N. (2015). Childhood hyperactivity/inattention and eating disturbances predict binge eating in adolescence. Psychological medicine, 45(12), 2511–2520. 10.1017/S0033291715000148

Steiger, J. H., & Lind, J. C. (1980, May 30). Statistically-based tests for the number of common factors [Paper presentation]. Annual Meeting of the Psychometric Society, Iowa City, IA, United States.

Svedlund, N. E., Norring, C., Ginsberg, Y., & von Hausswolff-Juhlin, Y. (2017). Symptoms of attention deficit hyperactivity disorder (ADHD) among adult eating disorder patients. BMC psychiatry, 17(1), 19. 10.1186/s12888-016-1093-1

Teague, S., Youssef, G. J., Macdonald, J. A., Sciberras, E., Shatte, A., Fuller-Tyszkiewicz, M.,…& SEED Lifecourse Sciences Theme Bant Sharyn Barker Sophie Booth Anna Capic Tanja Di Manno Laura Gulenc Alisha Le Bas Genevieve Letcher Primrose Lubotzky Claire Ann Opie Jessica O’Shea Melissa Tan Evelyn Williams Jo. (2018). Retention strategies in longitudinal cohort studies: a systematic review and meta-analysis. BMC medical research methodology, 18(1), 151. 10.1186/s12874-018-0586-7

Thomas, K. S., Cooper, K., & Jones, C. R. (2025). The intersection of autistic traits, ADHD traits, and gender diversity in disordered eating and drive for muscularity within the general population. Neurodiversity, 3, 27546330241308649. 10.1177/27546330241308649

Tistarelli, N., Fagnani, C., Troianiello, M., Stazi, M. A., & Adriani, W. (2020). The nature and nurture of ADHD and its comorbidities: A narrative review on twin studies. Neuroscience & Biobehavioral Reviews, 109, 63–77. 10.1016/j.neubiorev.2019.12.017

Tripp, G., & Wickens, J. R. (2008). Research review: dopamine transfer deficit: a neurobiological theory of altered reinforcement mechanisms in ADHD. Journal of child psychology and psychiatry, and allied disciplines, 49(7), 691–704. 10.1111/j.1469-7610.2007.01851.x

Trouton, A., Spinath, F. M., & Plomin, R. (2002). Twins early development study (TEDS): a multivariate, longitudinal genetic investigation of language, cognition and behavior problems in childhood. Twin Research and Human Genetics, 5(5), 444–448. 10.1375/twin.5.5.444

Tucker, L., & Lewis, C. (1973). A reliability coefcient for maximum likelihood factor analysis. Psychometrika, 38(1), 1–10. 10.1007/BF02291170

Verhulst, B., & Estabrook, R. (2012). Using genetic information to test causal relationships in cross-sectional data. Journal of theoretical politics, 24(3), 328–344. 10.1177/095162981243934

Waldman, I. D., & Gizer, I. R. (2006). The genetics of attention deficit hyperactivity disorder. Clinical psychology review, 26(4), 396–432. 10.1016/j.cpr.2006.01.007

You, Y., Oginni, O. A., Rijsdijk, F. V., Lim, K. X., Zavos, H. M., & McAdams, T. A. (2024). Exploring associations between ADHD symptoms and emotional problems from childhood to adulthood: shared aetiology or possible causal relationship?. Psychological Medicine, 54(15), 4231–4242. 10.1017/S0033291724002514

