## Supplemental Files for "Exploring the role of binge eating in the association between ADHD and BMI: A twin study"

**Supplementary materials**

| **Table S1**. The ADHD measurements at each stage | | | |
| --- | --- | --- | --- |
|  | ADHD | Hyperactivity-impulsivity | Inattention |
| Childhood | SDQ4^a^, SDQ7, SDQ9, Conners8^b^ | SDQ4, SDQ7, SDQ9, Conners8 | SDQ7, SDQ9, Conners8 |
| Adolescent | SDQ12, SDQ16, Conners12, Conners14 Conners16 | SDQ12, SDQ16, Conners12, Conners14 Conners16 | SDQ12, SDQ16, Conners12, Conners14 Conners16 |
| Adulthood | SDQ21^c^, Conners21, SDQ(T)21^d^ | SDQ21, Conners21, SDQ(T)21 | SDQ21, Conners21, SDQ(T)21 |

Note: ^a^SDQ4, Strengths and Difficulties Questionnaire (SDQ) at age 4;

^b^Conners8, Conners’ Rating Scales-Revised (Conners) at age 8;

^c^SDQ21, Strengths and Difficulties Questionnaire (SDQ) reported by parents at age 21;

^d^SDQ(T)21, Strengths and Difficulties Questionnaire (SDQ) reported by twins at age 21.

| **Table S2**. Univariate twin models for binge eating and BMI | | | | | | | | | |
| --- | --- | --- | --- | --- | --- | --- | --- | --- | --- |
| Model | -2LL | ep | AIC(weight) | *df* | CFI | TFI | RMSEA | *p* | Nested_*p* |
| 1 Binge eating_ACE | 42976 | 7 | 42990(0.27) | 9165 | 0.94 | 0.96 | 0.02 | 0 |  |
| **1.1 Binge eating_AE** | **42976** | **6** | **42988(0.73)** | **9166** | **0.94** | **0.97** | **0.02** | **0** | 1 |
| 2 BMI_ACE | 49065 | 7 | 49079(0.38) | 8187 | 0.997 | 0.998 | 0.01 | 0.07 |  |
| **2.1 BMI_AE** | **49066** | **6** | **49078(0.62)** | **8188** | **0.997** | **0.998** | **0.01** | **0.07** | **0.22** |

Note: ep, estimated parameters of the comparison model; -2LL, minus 2*log-likelihood of the comparison model; *df*, degrees in freedom of the comparison model.

The models in bold type are the best-fitting models.

Binge eating_AE/BMI_AE: we tested simplified models in which the C components were dropped. These simplified models were nested within the full ACE models.

*p*: *p* refers to the significance level of the chi-square test of model fit;

Nested_*p*: Nested_*p* indicates whether the simplified model fits significantly worse than the baseline model. When *p* > 0.05, it means there is no significant difference between the two models.

| **Table S3**. Estimates of shared genetic, dominant genetic, shared environmental, and non-shared environmental effects for binge eating and BMI | | | | | |
| --- | --- | --- | --- | --- | --- |
|  | ACE | | | AE | |
|  | h^2^ | c^2^ | e^2^ | h^2^ | e^2^ |
| Binge eating | 0.33(0.29,0.37) | 0(0,0.02) | 0.67(0.63,0.71) | 0.33(0.29,0.37) | 0.68(0.63,0.71) |
| BMI | 0.69(0.61,0.76) | 0.05(0,0.13) | 0.25(0.23,0.28) | 0.75(0.73,0.77) | 0.25(0.23,0.27) |

Note: h² represents the proportion of phenotypic variance attributed to additive genetic effects from the best-fitting models in each stage; c^2^ represents the proportion of phenotypic variance attributed to shared environmental effects from the best-fitting models in each stage; d² represents the proportion of phenotypic variance attributed to dominant genetic effects from the best-fitting models in each stage; e² represents the proportion of phenotypic variance attributed to non-shared environmental effects from the best-fitting models in each stage.

| **Table S4**. Descriptive statistics of observed variables | | | | | | |
| --- | --- | --- | --- | --- | --- | --- |
|  |  | Total | MZ | DZ | CorMZ | CorDZ |
|  |  | M(SD) | M(SD) | M(SD) |  |  |
| Childhood ADHD | SDQ4^a^ | 4.02(2.34) | 4.09(2.20) | 3.98(2.41) | 0.51** | -0.06** |
|  | SDQ7 | 3.64(2.58) | 3.63(2.51) | 3.63(2.61) | 0.60** | -0.19 |
|  | SDQ9 | 3.28(2.40) | 3.30(2.32) | 3.25(2.45) | 0.73** | 0.11** |
|  | Conners8^b^ | 11.38(9.57) | 11.42(9.50) | 11.32(9.57) | 0.84** | 0.38** |
| Adolescent ADHD | SDQ12 | 2.85(2.29) | 2.82(2.20) | 2.87(2.33) | 0.75** | 0.19** |
|  | SDQ16 | 2.28(2.00) | 2.16(1.87) | 2.35(2.06) | 0.72** | 0.21** |
|  | Conners12 | 9.97(8.77) | 9.85(8.63) | 10.01(8.83) | 0.85** | 0.40** |
|  | Conners14 | 8.71(8.48) | 8.25(7.93) | 8.98(8.77) | 0.85** | 0.38** |
|  | Conners16 | 6.91(7.56) | 6.25(7.03) | 7.26(7.81) | 0.77** | 0.40** |
| Adulthood ADHD | Conners21 | 6.57(7.20) | 5.94(6.73) | 6.91(7.43) | 0.73** | 0.35** |
|  | SDQ21^c^ | 1.96(2.00) | 1.76(1.83) | 2.06(2.09) | 0.59** | 0.17** |
|  | SDQ(T)21^d^ | 3.32(2.19) | 3.25(2,18) | 3.34(2.20) | 0.35** | 0.16** |
| Adulthood binge eating | Binge eating 21 | 3.75(2.57) | 3.69(2.51) | 3.78(2.60) | 0.36** | 0.10** |
| Adulthood BMI | BMI 26 | 24.74(5.22) | 24.58(5.10) | 24.82(5.26) | 0.74** | 0.38** |

Note: ^a^SDQ4, Strengths and Difficulties Questionnaire (SDQ) at age 4;

^b^Conners8, Conners’ Rating Scales-Revised (Conners) at age 8;

^c^SDQ21, Strengths and Difficulties Questionnaire (SDQ) reported by parents at age 21;

^d^SDQ(T)21, Strengths and Difficulties Questionnaire (SDQ) reported by twins at age 21.

| **Table S5**. Estimates of shared genetic, dominant genetic and specific environmental effects for each variable from the best-fitting models and factor correlations | | | | | | |
| --- | --- | --- | --- | --- | --- | --- |
| Developmental stages | ADHD | Binge eating | Adjusted BMI | h^2^ | d^2^ | e^2^ |
| 1 Childhood |  |  |  |  |  |  |
| 1.1 ADHD | 1 |  |  | 0.03(0.02,0.05) | 0.75(0.73,0,77) | 0.21(0.20,0.23) |
| 1.2 Binge eating | 0.06(0.03,0.08) | 1 |  | 0.33(0.29,0.37) | - | 0.67(0.63,0.71) |
| 1.3 Adjusted BMI | 0.11(0.08,0.14) | 0.24(0.22,0.26) | 1 | 0.73(0.70,0.76) | - | 0.27(0.24,0.30) |
| 2 Adolescence |  |  |  |  |  |  |
| 2.1 ADHD | 1 |  |  | 0.73(0.62,0.84) | 0.13(0.02,0.24) | 0.14(0.13,0.15) |
| 2.2 Binge eating | 0.10(0.08,0.13) | 1 |  | 0.33(0.29,0.37) | - | 0.67(0.63,0.71) |
| 2.3 Adjusted BMI | 0.17(0.14,0.20) | 0.24(0.22,0.27) | 1 | 0.63(0.59,0.67) | - | 0.37(0.33,0.41) |
| 3 Adulthood |  |  |  |  |  |  |
| 3.1 ADHD | 1 |  |  | 0.43(0.27,0.58) | 0.32(0.16,0.48) | 0.26(0.23,0.28) |
| 3.2 Binge eating | 0.25(0.22,0.28) | 1 |  | 0.32(0.28,0.36) |  | 0.67(0.64,0.72) |
| 3.3 Adjusted BMI | 0.18(0.14,0.21) | 0.24(0.22,0.26) | 1 | 0.27(0.21,0.33) |  | 0.73(0.67,0.79) |

Note: h² represents the proportion of phenotypic variance attributed to additive genetic effects from the best-fitting models in each stage; d² represents the proportion of phenotypic variance attributed to dominant genetic effects from the best-fitting models in each stage; e² represents the proportion of phenotypic variance attributed to non-shared environmental effects from the best-fitting models in each stage.

The heritability estimates for BMI from the best-fitting multivariate models were somewhat reduced because the previous-stage BMI was controlled for.

| **Table S6**. Parameter estimates (95% confidence intervals) in every stage | | | | | | | | | | | | | | |
| --- | --- | --- | --- | --- | --- | --- | --- | --- | --- | --- | --- | --- | --- | --- |
|  | a_11_ | a_12_ | a_13_ | a_22_ | a_23_ | a_33_ | e_11_ | e_12_ | e_13_ | e_22_ | e_23_ | e_33_ | d_11_ | Causal_2 on 3_ |
| Childhood_a | 0.03(0.02,0.05) | 0.16(0.06,0.26) | 0.63(0.42,0.74) | 0.17(0.07,0.28) | 0.11(0.00,0.31) | 0(0,0.43) | 0.21(0.20,0.23) | 0(0,0.003) | 0.00(0,0.01) | 0.67(0.63,0.71) | 0.01(0.00,0.01) | 0.26(0.23,0.29) | 0.75(0.72,0.77) | - |
| Childhood_b | 0.04(0.02,0.05) | 0.61(0.39,0.80) | 0.96(0.85,1.00) | 0.33(0.29,0.37) | 0.36(0.28,0.44) | 0.73(0.70,0.76) | 0.21(0.20,0.23) | - | - | 0.67(0.63,0.71) | - | 0.27(0.24,0.30) | 0.75(0.73,0.77) | 0.09(0.05,0.14) |
| Adolescence_a | 0.73(0.62,0.84) | 0.01(0.01,0.02) | 0.04(0.02,0.06) | 0.32(0.27,0.36) | 0.05(0.02,0.09) | 0.54(0.49,0.58) | 0.14(0.13,0.15) | 0(0,0.01) | 0(0,0.003) | 0.67(0.63,0.71) | 0.01(0.00,0.01) | 0.36(0.33,0.40) | 0.13(0.02,0.24) | - |
| Adolescence_b | 0.73(0.62,0.84) | 0.21(0.15,0.27) | 0.25(0.20,0.31) | 0.33(0.29,0.37) | 0.32(0.23,0.41) | 0.63(0.59,0.67) | 0.14(0.13,0.15) | - | - | 0.67(0.63,0.71) | - | 0.37(0.33,0.41) | 0.13(0.02,0.24) | 0.09(0.04,0.14) |
| Adulthood_a | 0.43(0.27,0.58) | 0.03(0.02,0.06) | 0.01(0.00,0.03) | 0.29(0.24,0.33) | 0.006(0,0.02) | 0.26(0.20,0.31) | 0.26(0.23,0.28) | 0.00(0.00,0.01) | 0(0,0.01) | 0.67(0.63,0.71) | 0(0,0.003) | 0.73(0.67,0.79) | 0.32(0.16,0.48) | - |
| Adulthood_b | 0.54(0.30,0.71) | - | - | 0.47(0.34,0.65) | - | 0.53(0.41,0.64) | 0.00(0.00,0.16) | - | - | 0.53(0.35,0.66) | - | 0.47(0.36,0.59) | 0.46(0.29,0.57) | - |

Note: In childhood and adolescence, the best-fitting models are Cholesky decomposition models (Childhood_a & Adolescence_a); In adulthood, the best -fitting models are either Cholesky decomposition models (Adulthood_a) or independent pathway models (Adulthood_b).

We also present the parameter estimates of both competing models when their fit was similar: Childhood_a, the Cholescky model in childhood; Childhood_b, the modified biometric mediation model in childhood; Adolescence_a, the Cholescky model in adolescence; Adolescence_b, the modified biometric mediation model in adolescence; Adulthood_a, the Cholescky model in adulthood; Adulthood_b, the independent pathway model in adulthood.

d_11_ represents the dominant genetic effect on ADHD;

Causal_2 on 3_ represents the standardized causal effect from binge eating to BMI;

If the best-fitting model is the Cholesky decomposition model, a_11_ represents the first genetic factor (A1) loading on ADHD, a_12_ represents the first genetic factor (A1) loading on binge eating, a_13_ represents the first genetic factor (A1) loading on BMI, a_22_ represents the second genetic factor (A2) loading on binge eating, a_23_ represents the second genetic factor (A2) loading on BMI, and a_33_ represents the third genetic factor (A3) loading on BMI; e_11_ represents the first non-shared environmental factor (E1) loading on ADHD, e_12_ represents the first non-shared environmental factor (E1) loading on binge eating, e_13_ represents the first non-shared environmental factor (E1) loading on BMI, e_22_ represents the second non-shared environmental factor (E2) loading on binge eating, e_23_ represents the second non-shared environmental factor (E2) loading on BMI, and e_33_ represents the third non-shared environmental factor (E3) loading on BMI (see Figure 2a).

If the best-fitting model is the Independent Pathway Model, a_11_ represents the additive genetic effect on inattention, a_22_ represents the additive genetic effect on binge eating, a_33_ represents the additive genetic effect on BMI, a_11_ represents the non-shared environmental effect on inattention, a_22_ represents the non-shared environmental effect on binge eating, and a_33_ represents the non-shared environmental effect on BMI (see Figure 2b).

If the best-fitting model is the modified biometric mediation model, which includes the correlations of genetic factors, a_12_ represents the genetic correlation between ADHD and binge eating, a_23_ represents the genetic correlation between binge eating and BMI, and a_13_ represents the genetic correlation between ADHD and BMI (see Figure 2c).

**Supplementary Text S1.**

**Phenotypic mediation models of ADHD subdomains, binge eating and BMI**

In the results of the phenotypic mediation models for hyperactivity-impulsivity, binge eating and BMI (see Table S7), the paths from hyperactivity-impulsivity to binge eating were significant in both adolescence (estimate = 0.79, 95% CI 0.35 to 1.25) and adulthood (estimate = 0.82, 95% CI 0.66 to 1.02), but not in childhood. Furthermore, the paths from hyperactivity-impulsivity to BMI were significant at all three stages, with estimates of 2.00 (95% CI 0.99 to 3.12) in childhood, 1.25 (95% CI 0.58 to 1.91) in adolescence, and 0.19 (95% CI 0.08 to 0.32) in adulthood. In addition, binge eating at age 21 mediated the relationship between hyperactivity-impulsivity and adulthood BMI in adolescence (estimate = 0.25, 95% CI 0.10 to 0.41) and early adulthood (estimate = 0.02, 95% CI 0.11 to 0.34), but not in childhood.

In the results of the phenotypic mediation models for inattention (see Table S8), the paths from inattention to binge eating were significant in both adolescence (estimate = 0.71, 95% CI 0.37 to 1.05) and adulthood (estimate = 0.07, 95% CI 0.04 to 0.11). In addition, after including the indirect paths, the direct paths from inattention to BMI were significant in childhood (estimate = 1.77, 95% CI 1.17 to 2.43) and adolescence (estimate = 1.06, 95% CI 0.59 to 1.53). However, the direct path from adulthood inattention to binge eating was not significant, although significant phenotypic correlations were found between the two traits (r = 0.21, 95% CI 0.18 to 0.24). Binge eating at age 21 mediated the associations between inattention and adult BMI in adolescence (estimate = 0.02, 95% CI 0.01 to 0.03) and adulthood (estimate = 0.05, 95% CI 0.02 to 0.07), but not in childhood. Therefore, the association between adulthood inattention and adulthood BMI was fully mediated by binge eating, whereas the association between adolescent inattention and adulthood BMI was partially mediated by binge eating.

| **Table S7**. The phenotypic mediation model of hyperactivity-impulsivity, binge eating, and BMI | | | | | |
| --- | --- | --- | --- | --- | --- |
| Path | Unstandardized estimates | 95% CIs | | *SE* | Standardized estimates |
|  |  | Lower | Upper |  |  |
| Direct effects |  |  |  |  |  |
| Childhood hyperactivity → BMI | 2.00 | 0.99 | 3.12 | 0.55 | 0.11 |
| Childhood hyperactivity→ Binge eating | 0.44 | -0.08 | 0.99 | 0.27 | 0.05 |
| Binge eating → BMI | 0.44 | 0.34 | 0.54 | 0.05 | 0.23 |
| Indirect effects |  |  |  |  |  |
| Hyperactivity→ Binge eating → BMI | 0.19 | -0.08 | 0.42 | 0.60 | 0.12 |
| Direct effects |  |  |  |  |  |
| Adolescent hyperactivity → BMI | 1.25 | 0.58 | 1.91 | 0.34 | 0.09 |
| Adolescent hyperactivity → Binge eating | 0.79 | 0.35 | 1.25 | 0.23 | 0.09 |
| Binge eating → BMI | 0.31 | 0.24 | 0.38 | 0.04 | 0.20 |
| Indirect effects |  |  |  |  |  |
| Hyperactivity → Binge eating → BMI | 0.25 | 0.10 | 0.41 | 0.08 | 0.02 |
| Direct effects |  |  |  |  |  |
| Adulthood hyperactivity → BMI | 0.19 | 0.08 | 0.32 | 0.06 | 0.05 |
| Adulthood hyperactivity → Binge eating | 0.82 | 0.66 | 1.02 | 0.09 | 0.25 |
| Binge eating → BMI | 0.07 | 0.03 | 0.10 | 0.02 | 0.06 |
| Indirect effects |  |  |  |  |  |
| Hyperactivity → Binge eating → BMI | 0.05 | 0.02 | 0.09 | 0.02 | 0.02 |

Note: CI, confidence interval; *SE*, standard error.

| **Table S8**. The phenotypic mediation model of inattention, binge eating, and BMI | | | | | |
| --- | --- | --- | --- | --- | --- |
| Path | Unstandardized estimates | 95% CIs | | *SE* | Standardized estimates |
|  |  | Lower | Upper |  |  |
| Direct effects |  |  |  |  |  |
| Childhood inattention → BMI | 1.77 | 1.17 | 2.43 | 0.31 | 0.15 |
| Childhood inattention → Binge eating | 0.08 | -0.27 | 0.43 | 0.18 | 0.01 |
| Binge eating → BMI | 0.44 | 0.34 | 0.55 | 0.05 | 0.24 |
| Indirect effects |  |  |  |  |  |
| Inattention→ Binge eating → BMI | 0.03 | -0.12 | 0.20 | 0.08 | 0.003 |
| Direct effects |  |  |  |  |  |
| Adolescent inattention → BMI | 1.06 | 0.59 | 1.53 | 0.24 | 0.09 |
| Adolescent inattention → Binge eating | 0.71 | 0.37 | 1.05 | 0.17 | 0.10 |
| Binge eating → BMI | 0.31 | 0.24 | 0.38 | 0.04 | 0.20 |
| Indirect effects |  |  |  |  |  |
| Inattention → Binge eating → BMI | 0.02 | 0.01 | 0.03 | 0.06 | 0.02 |
| Direct effects |  |  |  |  |  |
| Adulthood inattention → BMI | 0.06 | -0.03 | 0.14 | 0.05 | 0.02 |
| Adulthood inattention → Binge eating | 0.07 | 0.04 | 0.11 | 0.02 | 0.07 |
| Binge eating → BMI | 0.62 | 0.53 | 0.74 | 0.05 | 0.29 |
| Indirect effects |  |  |  |  |  |
| Inattention → Binge eating → BMI | 0.05 | 0.02 | 0.07 | 0.01 | 0.02 |

Note: CI, confidence interval; *SE*, standard error.

**Supplementary Text S2.**

**Biometric models of hyperactivity-impulsivity, binge eating and BMI**

First, we compared the results of the Cholesky decomposition model, the Independent pathway model, the biometric model and the modified biometric mediation model in childhood (See Table S9-S11). The AIC (264164) of the modified biometric mediation model and the Cholesky decomposition model were the same in childhood. This suggests no strong evidence to support either model. In the modified biometric mediation models, the genetic correlations between variables were significant. After controlling for genetic overlap, only the path from binge eating to BMI was significant. Regarding the Cholesky decomposition model, it involves no assumptions regarding causal relationships between variables. Based on the principle of parsimony, we selected the Cholesky decomposition model as the best-fitting model. This model revealed significant shared genetic components underlying the associations among hyperactivity-impulsivity, binge eating, and BMI. Specifically, the first genetic factor (A₁) loaded on all three traits, representing a common genetic factor influencing hyperactivity-impulsivity (a_11_ = 0.03, 95% CI 0.02 to 0.05), binge eating (a_12_ = 0.17, 95% CI 0.07, 0.28), and BMI (a_13_ = 0.59, 95% CI 0.37 to 0.73). The second genetic factor (A₂) influenced binge eating (a_22_ = 0.15, 95% CI 0.05 to 0.27) and BMI (a_23_ = 0.15, 95% CI 0.05 to 0.27), representing the residual shared genetic effects between the two after accounting for A₁. Apart from A₁ and A₂, no unique genetic effects (A₃) on BMI were observed. Regarding environmental influences, non-shared environmental components (E) accounted for 19% (95% CI 0.17 to 0.21), 67% (95% CI 0.63 to 0.71), and 26% (95% CI 0.23 to 0.29) of the variance in hyperactivity-impulsivity, binge eating, and BMI, respectively. However, the common non-shared environmental influences among the traits were close to 0. Therefore, the associations between childhood hyperactivity-impulsivity and adulthood binge eating and BMI were largely explained by common genetic factors, although it should be noted that the pathway from binge eating to BMI may also be significant.

We next compared the same set of models for hyperactivity–impulsivity, binge eating, and BMI using adolescent data. The AIC difference between the modified biometric mediation model (AIC = 267719) and the Cholesky decomposition model (AIC = 267720) was only 1. This suggests no strong evidence to support either model. In the modified biometric mediation models, genetic correlations were observed among the variables. After accounting for genetic overlap, only the paths from binge eating to BMI were significant. Regarding the Cholesky decomposition model, it makes no assumptions about causal relationships between variables. Based on the principle of parsimony, we selected the Cholesky decomposition model as the best-fitting model. This model included significant shared genetic components among ADHD, binge eating, and BMI. Specifically, the first genetic factor (A₁), which represents the common genetic effects influencing all three traits, significantly loaded on ADHD (a_11_ = 0.78, 95% CI 0.66 to 0.89), binge eating (a_12_ = 0.01, 95% CI 0.004 to 0.02), and BMI (childhood: a_13_ = 0.03, 95% CI 0.02 to 0.05). The second genetic factor (A₂) influenced binge eating (a_22_ = 0.32, 95% CI 0.28 to 0.36) and BMI (a_23_ = 0.05, 95% CI 0.02 to 0.09), representing the residual shared genetic effects between the two after accounting for A₁. Apart from A₁ and A₂, no unique genetic effects (A₃) on BMI were observed in childhood, whereas unique genetic effects on BMI were still observed in adolescence (a_33_ = 0.54, 95% CI 0.49 to 0.59). Regarding environmental influences, common non-shared environmental influences among the traits were small and non-significant. This indicates that the non-shared environmental components (E) on the variables were independent of each other: 11% (95% CI 0.10 to 0.12), 67% (95% CI 0.63 to 0.71), and 37% (95% CI 0.33 to 0.41) of the variance in ADHD, binge eating, and BMI in childhood, respectively. Therefore, the associations between adolescent ADHD, binge eating, and BMI were largely explained by common genetic factors, although it should be noted that the pathway from binge eating to BMI may also be significant.

In adulthood, the Cholesky decomposition model with an AIC of 187422 was selected as the best-fitting model. However, its AIC was only 1 point lower than that of the independent pathway model and the biometric mediation model. Therefore, we acknowledge that there is no strong evidence favoring any one of the three models, and it remains possible that the independent pathway model or the biometric mediation model may represent the underlying structure. Within the Cholescky decomposition model, specifically, the first genetic factor (A₁), which represents the common genetic effects influencing all three traits, significantly loaded on hyperactivity-impulsivity (a_11_ = 0.28, 95% CI 0.09 to 0.46), binge eating (a_12_ = 0.02, 95% CI 0.03 to 0.06), and BMI (a_13_ = 0.02, 95% CI 0.03 to 0.06). The second genetic factor (A₂) significantly influenced binge eating (a_22_ = 0.29, 95% CI 0.20 to 0.33), but not BMI, indicating that there were no residual shared genetic effects between the two after accounting for A₁. In addition, there was also a significant genetic effect (A₃) only influencing BMI (a_33_ = 0.25, 95% CI 0.18 to 0.31). Regarding environmental influences, non-shared environmental components (E) accounted for 19% (95% CI 0.17 to 0.21), 67% (95% CI 0.63 to 0.71), and 78% (95% CI 0.76 to 0.80) of the variance in hyperactivity-impulsivity, binge eating, and BMI, respectively. However, the paths that indicated common non-shared environmental influences among the traits were small and non-significant. Therefore, the associations between childhood hyperactivity-impulsivity and adulthood binge eating and BMI were largely explained by common genetic factors.

**Biometric models of inattention, binge eating and BMI**

First, we examined the genetic structure among childhood inattention, binge eating, and BMI (See Table S12-S14). The AIC difference between the modified biometric mediation model (AIC = 206635) and the Cholesky decomposition model (AIC = 206637) was only 2 in childhood. This suggests no strong evidence to support either model. In the modified biometric mediation models, genetic correlations were observed among the variables. After accounting for genetic overlap, only the paths from binge eating to BMI were significant. Regarding the Cholesky decomposition model, it makes no assumptions about causal relationships between variables. Based on the principle of parsimony, the Cholesky decomposition model was selected as the best-fitting model. The Cholesky decomposition model revealed significant shared genetic influences among the three traits. Specifically, the first genetic factor (A₁), which represents the common genetic effects influencing all three traits, significantly loaded on inattention (a_11_ = 0.03, 95% CI 0.01 to 0.04), binge eating (a_12_ = 0.13, 95% CI 0.03 to 0.25), and BMI (a_13_ = 0.66, 95% CI 0.44 to 0.75). The second genetic factor (A₂) significantly influenced binge eating (a_22_ = 0.20, 95% CI 0.08 to 0.31), but not BMI, indicating that there were no residual shared genetic effects between the two after accounting for A₁. Apart from A₁ and A₂, no unique genetic effects (A₃) on BMI were observed. Regarding environmental influences, the non-shared environmental components (E) accounted for 27% (95% CI 0.25 to 0.30), 67% (95% CI 0.63 to 0.71), and 26% (95% CI 0.23 to 0.29) of the variance in inattention, binge eating, and BMI, respectively. However, the paths that indicated common non-shared environmental influences among the traits were small and non-significant. Therefore, the associations between childhood inattention and adulthood binge eating and BMI were largely explained by common genetic factors, although it should be noted that the pathway from binge eating to BMI may also be significant.

In adolescence and adulthood, the Cholesky decomposition model (AICs = 273,637 and 182,795, respectively) and the independent pathway model with the same AIC values demonstrated the best fit to the data (see Tables S8-S10). Specifically, in the Cholescky decomposition model, a common genetic factor (A₁) that significantly influenced ADHD (adolescence: a_11_ = 0.30, 95% CI 0.17 to 0.42; adulthood: a_11_ = 0.13, 95% CI 0.03 to 0.30), binge eating (adolescence: a_12_ = 0.03, 95% CI 0.01 to 0.06; adulthood: a_12_ = 0.11, 95% CI 0.04 to 0.35), and BMI (adolescence: a_13_ = 0.08, 95% CI 0.04 to 0.15; adulthood: a_13_ = 0.02, 95% CI 0.00 to 0.16) was found. In addition, another common genetic factor (A₂) was identified, representing the residual shared genetic effects between binge eating (adolescence: a_22_ = 0.30, 95% CI 0.25 to 0.34; adulthood: a_22_ = 0.33, 95% CI 0.29 to 0.37) and BMI (adolescence: a_23_ = 0.03, 95% CI 0.01 to 0.07; adulthood: a_23_ = 0.002, 95% CI 0.00 to 0.30) after accounting for A₁. Finally, there was also a significant genetic effect (A₃) that influenced only BMI (adolescence: a_33_ = 0.51, 95% CI 0.44 to 0.57; adulthood: a_33_ = 0.27, 95% CI 0.21 to 0.33). Regarding environmental influences, the non-shared environmental components (E) accounted for 18% (95% CI 0.17 to 0.20), 67% (95% CI 0.63 to 0.71), and 36% (95% CI 0.33 to 0.40) of the variance in ADHD, binge eating, and BMI respectively in adolescence; it accounted for 28% (95% CI 0.26 to 0.31), 67% (95% CI 0.63 to 0.71), and 73% (95% CI 0.67 to 0.79) of the variance in ADHD, binge eating, and BMI respectively in adulthood. However, the common non-shared environmental influences among the traits were small and non-significant. In the independent pathway model, a set of common genetic and environmental factors influenced all three traits. The results indicated that the common genetic factor accounted for 34% (95% CI 0.19 to 0.40), 49% (95% CI 0.36 to 0.63), and 98% (95% CI 0.88 to 0.997) of the variance in inattention, binge eating, and BMI respectively in adolescence. The common non-shared environmental factor accounted for 13% (95% CI 0.03–0.30), 33% (95% CI 0.29 to 0.37) and 27% (95% CI 0.21 to 0.33) of the variance in ADHD, binge eating and BMI. In total, the best models for adulthood, after accounting for shared genetic and environmental factors, did not include any paths between the phenotypes.

| **Table S9**. Estimates of shared genetic, dominant genetic and non-shared environmental effects for each variable from the best-fitting models and factor correlations _hyperactivity-impulsivity | | | | | | |
| --- | --- | --- | --- | --- | --- | --- |
| Developmental stages | Hyperactivity-impulsivity | Binge eating | Adjusted BMI | h^2^ | d^2^ | e^2^ |
| 1 Childhood |  |  |  |  |  |  |
| 1.1 Hyperactivity-impulsivity | 1 |  |  | 0.03(0.02,0.05) | 0.78(0.76,0.80) | 0.19(0.17,0.21) |
| 1.2 Binge eating | 0.06(0.04,0.09) | 1 |  | 0.33(0.29,0.37) | - | 0.67(0.63,0.71) |
| 1.3 Adjusted BMI | 0.11(0.08,0.13) | 0.24(0.22,0.26) | 1 | 0.73(0.70,0.76) | - | 0.27(0.24,0.30) |
| 2 Adolescence |  |  |  |  |  |  |
| 2.1 Hyperactivity-impulsivity | 1 |  |  | 0.78(0.66,0.89) | 0.11(0,0.23) | 0.11(0.10,0.12) |
| 2.2 Binge eating | 0.08(0.05,0.11) | 1 |  | 0.33(0.29,0.37) | - | 0.67(0.63,0.71) |
| 2.3 Adjusted BMI | 0.14(0.11,0.17) | 0.24(0.22,0.26) | 1 | 0.63(0.59,0.66) | - | 0.37(0.34,0.41) |
| 3 Adulthood |  |  |  |  |  |  |
| 3.1 Hyperactivity-impulsivity | 1 |  |  | 0.28(0.09,0.46) | 0.45(0.27,0.63) | 0.28(0.25,0.31) |
| 3.2 Binge eating | 0.13(0.10,0.15) | 1 |  | 0.33(0.29,0.37) | - | 0.67(0.63,0.71) |
| 3.3 Adjusted BMI | 0.10(0.07,0.14) | 0.24(0.22,0.26) | 1 | 0.27(0.21,0.33) | - | 0.73(0.67,0.79) |

Note: h² represents the proportion of phenotypic variance attributed to additive genetic effects from the best-fitting models in each stage; d² represents the proportion of phenotypic variance attributed to dominant genetic effects from the best-fitting models in each stage; e² represents the proportion of phenotypic variance attributed to non-shared environmental effects from the best-fitting models in each stage.

The heritability estimates for BMI from the best-fitting multivariate models were somewhat reduced because the previous-stage BMI was controlled for.

| **Table S10**. Fit statistics for Cholesky decomposition models, Independent Pathway Models and Adjusted biometric mediation models at each stage_hyperactivity-impulsivity | | | | | | | | | |
| --- | --- | --- | --- | --- | --- | --- | --- | --- | --- |
| Model | -2LL | ep | AIC (weight) | df | CFI | TFI | RMSEA | *p* | Nested_*p* |
| 1. Phenotypic mediation model |  |  |  |  |  |  |  |  |  |
| Phenotypic mediation model_childhood | 30414 | 21 | 30457 | 6 | 0.996 | 0.991 | 0.027 | 0.04 |  |
| Phenotypic mediation model_adolescence | 60415 | 26 | 60467 | 9 | 0.983 | 0.96 | 0.066 | 0 |  |
| Phenotypic mediation models_adulthood | 85733 | 17 | 85767 | 3 | 1 | 1 | 0 | 0 |  |
| 2. Childhood |  |  |  |  |  |  |  |  |  |
| **2.1 Cholesky decomposition model** | **264124** | **37** | **264199(0.36)** | **63324** | **0.99** | **0.99** | **0.01** | **0** | **-** |
| 2.2 Independent pathway model | 264129 | 37 | 264203(0.05) | 63324 | 0.99 | 0.99 | 0.01 | 0 | - |
| 2.3 Biometric mediation model | 264162 | 34 | 264230(0.00) | 63327 | 0.99 | 0.99 | 0.01 | 0 | - |
| 2.3 Modified biometric mediation model | 264124 | 37 | 264198(0.59) | 63324 | 0.99 | 0.99 | 0.01 | 0 | - |
| 2.4 Modified biometric mediation model_nested | 264127 | 35 | 264197 | 63326 | 0.99 | 0.99 | 0.01 | 0 | 0.21 |
| 3. Adolescence |  |  |  |  |  |  |  |  |  |
| **3.1 Cholesky decomposition model** | **267720** | **41** | **267802(0.38)** | **63169** | **0.93** | **0.93** | **0.03** | **0** | **-** |
| 3.2 Independent pathway model | 267778 | 41 | 267861(0.00) | 63169 | 0.93 | 0.93 | 0.03 | 0 | - |
| 3.3 Biometric mediation model | 267748 | 38 | 267824(0.00) | 63172 | 0.93 | 0.93 | 0.03 | 0 | - |
| 3.4 Modified biometric mediation model | 267719 | 41 | 267801(0.62) | 63169 | 0.93 | 0.93 | 0.03 | 0 | - |
| 3.5 Modified biometric mediation model_nested | 267722 | 39 | 267800 | 63171 | 0.93 | 0.93 | 0.03 | 0 | 0.19 |
| 4 Adulthood |  |  |  |  |  |  |  |  |  |
| **4.1 Cholesky decomposition models** | **187422** | **30** | **187482(0.45)** | **45146** | **0.94** | **0.95** | **0.02** | **0** | **-** |
| 4.2 Independent pathway models | 187423 | 30 | 187483(0.27) | 45146 | 0.94 | 0.95 | 0.02 | 0 | - |
| 4.3 Biometric mediation model | 187436 | 27 | 187490(0.01) | 45149 | 0.94 | 0.95 | 0.02 | 0 | - |
| 4.3 Modified biometric mediation models | 187423 | 30 | 187483(0.27) | 45146 | 0.94 | 0.95 | 0.02 | 0 | - |

Note: ep, estimated parameters of the comparison model; -2LL, minus 2*log-likelihood of the comparison model; *df*, degrees in freedom of the comparison model.

The models in bold type are the best-fitting models.

Adjusted biometric mediation model_nested: we tested a simplified model in which the paths from ADHD to binge eating and ADHD to BMI were dropped. This simplified model was nested within the full Adjusted biometric mediation model

*p*: *p* refers to the significance level of the chi-square test of model fit;

Nested_*p*: Nested_p indicates whether the simplified model fits significantly worse than the baseline model. When *p* > 0.05, it means there is no significant difference between the two models.

**Table S11**. Parameter estimates (95% confidence intervals) at each stage_hyperactivity-impulsivity

|  | a_11_ | a_12_ | a_13_ | a_22_ | a_23_ | a_33_ | e_11_ | e_12_ | e_13_ | e_22_ | e_23_ | e_33_ | d_11_ | Causal_2 on 3_ |
| --- | --- | --- | --- | --- | --- | --- | --- | --- | --- | --- | --- | --- | --- | --- |
| Childhood_a | 0.03(0.02,0.05) | 0.17(0.07,0.28) | 0.59(0.37, 0.73) | 0.15(0.05,0.27) | 0.15(0.05,0.27) | 0.00(0.00,0.52) | 0.19(0.17,0.21) | 0(0,0.01) | 0(0,00,0.01) | 0.67(0.63,0.71) | 0.01(0,0.01) | 0.26(0.23,0.29) | 0.78(0.76,0.80) |  |
| Childhood_b | 0.03(0.02,0.05) | 0.63(0.39,0.83) | 0.95(-0.61,1.00) | 0.33(0.29,0.37) | 0.36(0.28,0.44) | 0.73(0.70,0.76) | 0.19(0.17,0.21) | - | - | 0.67(0.63,0.71) | - | 0.27(0.24,0.30) | 0.78(0.76,0.80) | 0.09(0.05,0.14) |
| Adolescence_a | 0.78(0.66,0.89) | 0.01(0.004,0.02) | 0.03(0.02,0.05) | 0.32(0.28,0.36) | 0.05(0.02,0.09) | 0.54(0.49,0.59) | 0.11(0.10,0.12) | 0(0,0.01) | 0(0,0.01) | 0.67(0.63,0.71) | 0.01(0,0.01) | 0.37(0.33,0.41) | 0.11(0,0.23) | - |
| Adolescence_b | 0.78(0.66,0.89) | 0.17(0.11,0.23) | 0.21(0.16,0.27) | 0.33(0.29,0.37) | 0.32(0.24,0.41) | 0.63(0.59,0.66) | 0.11(0.10,0.12) | - | - | 0.67(0.63,0.71) | - | 0.37(0.34,0.41) | 0.11(0,0.23) | 0.09(0.04,0.14) |
| Adulthood_a | 0.28(0.09,0.46) | 0.04(0.02,0.13) | 0.02(0.003,0.06) | 0.29(0.20,0.33) | 0.003(0,0.02) | 0.25(0.18,0.31) | 0.28(0.25,0.31) | 0.002(0,0.01) | 0(0,0.01) | 0.67(0.63,0.71) | 0(0,0.003) | 0.73(0.67,0.79) | 0.45(0.28,0.63) | - |

Note: In childhood, adolescence and adulthood, the best-fitting models are Cholesky decomposition models (Childhood_a, Adolescence_a & Adulthood_a).

We also present the parameter estimates of both competing models when their fit was similar: Childhood_a, the Cholescky decomposition model in childhood; Childhood_b, the modified biometric mediation model in childhood; Adolescence_a, the Cholescky decomposition model in adolescence; Adolescence_b, the modified biometric mediation model in adolescence; Adulthood_a, the Cholescky decomposition model in adulthood.

d_11_ represents the dominant genetic effect on hyperactivity-impulsivity;

Causal_2 on 3_ represents the standardized causal effect from binge eating to BMI;

If the best-fitting model is the Cholesky decomposition model, a_11_ represents the first genetic factor (A1) loading on hyperactivity-impulsivity, a_12_ represents the first genetic factor (A1) loading on binge eating, a_13_ represents the first genetic factor (A1) loading on BMI, a_22_ represents the second genetic factor (A2) loading on binge eating, a_23_ represents the second genetic factor (A2) loading on BMI, and a_33_ represents the third genetic factor (A3) loading on BMI; e_11_ represents the first non-shared environmental factor (E1) loading on hyperactivity-impulsivity, e_12_ represents the first non-shared environmental factor (E1) loading on binge eating, e_13_ represents the first non-shared environmental factor (E1) loading on BMI, e_22_ represents the second non-shared environmental factor (E2) loading on binge eating, e_23_ represents the second non-shared environmental factor (E2) loading on BMI, and e_33_ represents the third non-shared environmental factor (E3) loading on BMI (see Figure 2a).

If the best-fitting model is the Adjusted biometric mediation model, which includes the correlations of genetic factors, a_12_ represents the genetic correlation between hyperactivity-impulsivity and binge eating, a_23_ represents the genetic correlation between binge eating and BMI, and a_13_ represents the genetic correlation between hyperactivity-impulsivity and BMI (see Figure 2c).

| **Table S12**. Estimates of shared genetic, dominant genetic and non-shared environmental effects for each variable from the best-fitting moels and factor correlations _inattention | | | | | | |
| --- | --- | --- | --- | --- | --- | --- |
| Developmental stages | Inattention | Binge eating | Adjusted BMI | h^2^ | d^2^ | e^2^ |
| 1 Childhood |  |  |  |  |  |  |
| 1.1 Inattention | 1 |  |  | 0.03(0.01,0.04) | 0.70(0.67,0.73) | 0.27(0.25,0.30) |
| 1.2 Binge eating | 0.05(0.02,0.08) | 1 |  | 0.33(0.29,0.37) | - | 0.67(0.63,0.71) |
| 1.3 Adjusted BMI | 0.15(0.12,0.18) | 0.24(0.22,0.26) | 1 | 0.73(0.70,0.76) | - | 0.27(0.24,0.30) |
| 2 Adolescence |  |  |  |  |  |  |
| 2.1 Inattention | 1 |  |  | 0.34(0.19,0.40) | 0.66(0.60,0.75) | 0.001(0,0.07) |
| 2.2 Binge eating | 0.11(0.08,0.13) | 1 |  | 0.49(0.36,0.63) | - | 0.51(0.37,0.64) |
| 2.3 Adjusted BMI | 0.17(0.14,0.20) | 0.24(0.22,0.27) | 1 | 0.98(0.88,0.996) | - | 0.02(0.003,0.12) |
| 3 Adulthood |  |  |  |  |  |  |
| 3.1 Inattention | 1 |  |  | 0.13(0.03,0.30) | 0.58(0.42,0.69) | 0.28(0.26,0.31) |
| 3.2 Binge eating | 0.21(0.18,0.24) | 1 |  | 0.33(0.29,0.37) | - | 0.67(0.63,0.71) |
| 3.3 Adjusted BMI | 0.14(0.10,0.18) | 0.24(0.21,0.27) | 1 | 0.27(0.21,0.33) | - | 0.73(0.67,0.79) |

Note: h² represents the proportion of phenotypic variance attributed to additive genetic effects from the best-fitting models in each stage; d² represents the proportion of phenotypic variance attributed to dominant genetic effects from the best-fitting models in each stage; e² represents the proportion of phenotypic variance attributed to non-shared environmental effects from the best-fitting models in each stage.

The heritability estimates for BMI from the best-fitting multivariate models were somewhat reduced because the previous-stage BMI was controlled for.

| **Table S13**. Fit statistics for Cholesky decomposition models, Independent Pathway Models and Adjusted biometric mediation models at each stage_inattention | | | | | | | | | |
| --- | --- | --- | --- | --- | --- | --- | --- | --- | --- |
| Model | -2LL | ep | AIC (weight) | df | CFI | TFI | RMSEA | *p* | Nested_*p* |
| 1. Phenotypic mediation model |  |  |  |  |  |  |  |  |  |
| Phenotypic mediation model_childhood | 24002 | 21 | 24044 | 6 | 1 | 1 | 0.01 | 0.35 |  |
| Phenotypic mediation model_adolescence | 65664 | 26 | 65716 | 9 | 0.99 | 0.97 | 0.06 | 0 |  |
| Phenotypic mediation models_adulthood | 90754 | 17 | 90788 | 3 | 0.999 | 0.999 | 0.01 | 0.15 |  |
| 2. Childhood |  |  |  |  |  |  |  |  |  |
| **2.1 Cholesky decomposition model** | **206637** | **30** | **206697(0.27)** | **48179** | **0.98** | **0.98** | **0.01** | **0** | **-** |
| 2.2 Independent pathway model | 206656 | 30 | 206716(0.00) | 48179 | 0.98 | 0.98 | 0.01 | 0 | - |
| 2.3 Biometric mediation model | 206671 | 27 | 206725(0.00) | 48182 | 0.98 | 0.98 | 0.01 | 0 | - |
| 2.4 Modified biometric mediation model | 206635 | 30 | 206695(0.73) | 48179 | 0.98 | 0.98 | 0.01 | 0 | - |
| 2.5 Modified biometric mediation model_nested | 206638 | 28 | 206694 | 48181 | 0.98 | 0.98 | 0.01 | 0 | 0.19 |
| 3. Adolescence |  |  |  |  |  |  |  |  |  |
| **3.1 Cholesky decomposition models** | **273637** | **41** | **273719(0.42)** | **63157** | **0.91** | **0.91** | **0.03** | **0** | **-** |
| **3.2 Independent pathway model** | **273637** | **41** | **273719(0.42)** | **63157** | **0.91** | **0.91** | **0.03** | **0** | **-** |
| 3.3 Biometric mediation model | 273661 | 38 | 273737(0.00) | 63160 | 0.91 | 0.91 | 0.03 | 0 | - |
| 3.3 Modified biometric mediation models | 273684 | 41 | 273721(0.16) | 63157 | 0.91 | 0.91 | 0.03 | 0 | - |
| 4. Adulthood |  |  |  |  |  |  |  |  |  |
| **4.1 Cholesky decomposition models** | **182795** | **30** | **182855(0.47)** | **45158** | **0.96** | **0.96** | **0.02** | **0** | **-** |
| **4.2 Independent pathway models** | **182795** | **30** | **182855(0.47)** | **45158** | **0.96** | **0.96** | **0.02** | **0** | **-** |
| 4.3 Biometric mediation models | 182808 | 27 | 182862(0.01) | 45161 | 0.96 | 0.96 | 0.02 | 0 | - |
| 4.4 Modified biometric mediation models | 182800 | 30 | 182860(0.04) | 45158 | 0.96 | 0.96 | 0.02 | 0 | - |

Note: ep, estimated parameters of the comparison model; -2LL, minus 2*log-likelihood of the comparison model; *df*, degrees in freedom of the comparison model.

The models in bold type are the best-fitting models.

Adjusted biometric mediation model_nested: we tested a simplified model in which the paths from ADHD to binge eating and ADHD to BMI were dropped. This simplified model was nested within the full Adjusted biometric mediation model.

*p*: *p* refers to the significance level of the chi-square test of model fit;

Nested_*p*: Nested_p indicates whether the simplified model fits significantly worse than the baseline model. When *p* > 0.05, it means there is no significant difference between the two models.

| **Table S14**. Parameter estimates (95% confidence intervals) at each stage_inattention | | | | | | | | | | | | | | |
| --- | --- | --- | --- | --- | --- | --- | --- | --- | --- | --- | --- | --- | --- | --- |
|  | a_11_ | a_12_ | a_13_ | a_22_ | a_23_ | a_33_ | e_11_ | e_12_ | e_13_ | e_22_ | e_23_ | e_33_ | d_11_ | Causal_2 on 3_ |
| Childhood_a | 0.03(0.01,0.04) | 0.13(0.03,0.25) | 0.66(0.44,0.75) | 0.20(0.08,0.31) | 0.07(0,0.29) | 0(0,0.35) | 0.27(0.25,0.30) | 0(0,0.003) | 0(0,0.005) | 0.67(0.63,0.71) | 0.01(0.00,0.01) | 0.26(0.23,0.29) | 0.70(0.67,0.73) |  |
| Childhood_b | 0.03(0.02,0.04) | 0.57(0.30,0.79) | 0.97(0.86,1.00) | 0.33(0.29,0.37) | 0.36(0.28,0.44) | 0.73(0.70,0.76) | 0.27(0.25,0.30) | - | - | 0.67(0.63,0.71) | - | 0.27(0.24,0.30) | 0.70(0.67,0.72) | 0.10(0.05,0.14) |
| Adolecence_a | 0.30(0.17,0.42) | 0.03(0.01,0.06) | 0.08(0.04,0.15) | 0.30(0.25,0.34) | 0.03(0.01,0.07) | 0.51(0.44,0.57) | 0.18(0.17,0.20) | 0.001(0,0.01) | 0(0,0) | 0.67(0.63,0.71) | 0.01(0.00,0.01) | 0.36(0.33,0.40) | 0.52(0.39,0.65) |  |
| Adolescence_b | 0.34(0.19,0.40) | - | - | 0.49(0.36,0.63) | - | 0.98(0.88,1.00) | 0.001(0,0.07) | - | - | 0.51(0.37,0.64) | - | 0.02(0.00,0.12) | 0.66(0.60,0.75) |  |
| Adulthood_a | 0.13(0.03,0.30) | 0.11(0.04,0.35) | 0.02(0,0.16) | 0.33(0.29,0.37) | 0.002(0,0.30) | 0.27(0.21,0.33) | 0.28(0.26,0.31) | 0.005(0,0.01) | 0.001(0,0.01) | 0.67(0.63,0.71) | 0(0,0.002) | 0.73(0.67,0.79) | 0.58(0.42,0.69) |  |
| Adulthood_b | 0.15(0.03,0.37) | - | - | 0.56(0.45,0.64) | - | 0.53(0.42,0.64) | 0.19(0.07,0.31) | - | - | 0.44(0.36,0.55) | - | 0.47(0.36,0.58) | 0.66(0.51,0.77) |  |

Note: In childhood, the best-fitting models are Cholesky decomposition models (Childhood_a); In adolescence and adulthood, the best -fitting models are either Cholesky decomposition models (Adolescence_a & Adulthood_a) or independent pathway models (Adolescence_b & Adulthood_b).

We also present the parameter estimates of both competing models when their fit was similar: Childhood_a, the Cholesky decomposition model in childhood; Childhood_b, the modified biometric mediation model in childhood; Adolescence_a, the Cholescky decomposition model in adolescence; Adolescence_b, the independent pathway model in adolescence; Adulthood_a, the Cholesky decomposition model in adulthood; Adulthood_b, the independent pathway model in adulthood.

d_11_ represents the dominant genetic effect on inattention;

Causal_2 on 3_ represents the standardized causal effect from binge eating to BMI;

If the best-fitting model is the Cholesky decomposition model, a_11_ represents the first genetic factor (A1) loading on inattention, a_12_ represents the first genetic factor (A1) loading on binge eating, a_13_ represents the first genetic factor (A1) loading on BMI, a_22_ represents the second genetic factor (A2) loading on binge eating, a_23_ represents the second genetic factor (A2) loading on BMI, and a_33_ represents the third genetic factor (A3) loading on BMI; e_11_ represents the first non-shared environmental factor (E1) loading on ADHD, e_12_ represents the first non-shared environmental factor (E1) loading on binge eating, e_13_ represents the first non-shared environmental factor (E1) loading on BMI, e_22_ represents the second non-shared environmental factor (E2) loading on binge eating, e_23_ represents the second non-shared environmental factor (E2) loading on BMI, and e_33_ represents the third non-shared environmental factor (E3) loading on BMI (see Figure 2a).

If the best-fitting model is the Independent Pathway Model, a_11_ represents the additive genetic effect on inattention, a_22_ represents the additive genetic effect on binge eating, a_33_ represents the additive genetic effect on BMI, a_11_ represents the non-shared environmental effect on inattention, a_22_ represents the non-shared environmental effect on binge eating, and a_33_ represents the non-shared environmental effect on BMI (see Figure 2b).

If the best-fitting model is the Adjusted biometric mediation model, which includes the correlations of genetic factors, a_12_ represents the genetic correlation between inattention and binge eating, a_23_ represents the genetic correlation between binge eating and BMI, and a_13_ represents the genetic correlation between inattention and BMI (see Figure 2c).

**Sensitive analysis**

The two subdomains of ADHD are strongly correlated, and this correlation may confound the evaluation of subdomain models. However, to preserve statistical power, we did not control for symptoms of the other subdomain in the ADHD subdomain models. As a sensitivity analysis, we repeated all analyses for childhood—the developmental stage with the largest sample size—while controlling for the other subdomain. The results remained consistent with our primary findings: the Cholesky decomposition model provided the best fit for both the inattention and hyperactivity-impulsivity models in childhood (see Table S15). Furthermore, these Cholesky decomposition models revealed significant common genetic components, but no significant non-shared environmental components (see Table S16), indicating that the associations among the three traits are primarily explained by genetic correlations.

| **Table S15.** Fit statistics for Cholesky models, independent pathway models and biometric mediation models at childhood_after controlling for another subdomain | | | | | | | | | |
| --- | --- | --- | --- | --- | --- | --- | --- | --- | --- |
| Model | -2LL | ep | AIC (weight) | *df* | CFI | TFI | RMSEA | *p* | Nested_*p* |
| 1 Childhood hyperactivity-impulsivity |  |  |  |  |  |  |  |  |  |
| **1,1 Cholesky models** | **244367** | **37** | **244442** | **59721** | **0.98** | **0.98** | **0.01** | **0** | - |
| 1.2 Independent pathway models | 244370 | 37 | 244444 | 59721 | 0.98 | 0.98 | 0.01 | 0 | - |
| 1.3 Biometric mediation models | 244402 | 34 | 244470 | 59724 | 0.97 | 0.98 | 0.01 | 0 | - |
| 1.4 Modified biometric mediation models | 244368 | 37 | 244442 | 59721 | 0.98 | 0.98 | 0.01 | 0 | - |
| 2 Childhood inattention |  |  |  |  |  |  |  |  |  |
| **2.1 Cholesky models** | **197524** | **30** | **197585** | **48145** | **0.97** | **0.97** | **0.01** | **0** | - |
| 2.2 Independent pathway models | 197525 | 30 | 197585 | 48145 | 0.97 | 0.97 | 0.01 | 0 | - |
| 2.3 Biometric mediation model | 197556 | 27 | 197610 | 48148 | 0.96 | 0.97 | 0.01 | 0 | - |
| 2.4 Adjusted biometric mediation models | 197524 | 30 | 197584 | 48145 | 0.97 | 0.97 | 0.01 | 0 | - |
| 2.5 Adjusted biometric mediation model_nested | 197524 | 28 | 197581 | 48147 | 0.97 | 0.97 | 0.01 | 0 | _ |

Note: ep, estimated parameters of the comparison model; -2LL, minus 2*log-likelihood of the comparison model; *df*, degrees in freedom of the comparison model.

The models in bold type are the best-fitting models.

Adjusted biometric mediation model_nested: we tested a simplified model in which the paths from ADHD to binge eating and ADHD to BMI were dropped. This simplified model was nested within the full Adjusted biometric mediation model

*p*: *p* refers to the significance level of the chi-square test of model fit;

Nested_*p*: Nested_p indicates whether the simplified model fits significantly worse than the baseline model. When *p* > 0.05, it means there is no significant difference between the two models.

| **Table S16.** Parameter estimates (95% confidence intervals) in childhood _after controlling for another subdomain | | | | | | | | | | | | | |
| --- | --- | --- | --- | --- | --- | --- | --- | --- | --- | --- | --- | --- | --- |
|  | a_11_ | a_12_ | a_13_ | a_22_ | a_23_ | a_33_ | e_11_ | e_12_ | e_13_ | e_22_ | e_23_ | e_33_ | d_11_ |
| Hyperactivity | 0.01(0.00,0.03) | 0.19(0.02,0.33) | 0.56(0.17,0.75) | 0.14(0.01,0.31) | 0.17(0.00,0.56) | 0.00(0.00,0.63) | 0.19(0.17,0.22) | 0.00(0.00,0.01) | 0.00(0.00,0.01) | 0.67(0.63,0.71) | 0.01(0.00,0.01) | 0.26(0.23,0.29) | 0.80(0.77,0.82) |
| Inattention | 0.01(0.00,0.02) | 0.08(0.00,0.28) | 0.71(0.35,076) | 0.25(0.05,0.35) | 0.02(0.00,0.38) | 0.00(0.00,0.49) | 0.37(0.33,0.41) | 0.00(0.00,0.00) | 0.00(0.00,0.00) | 0.67(0.63,0.71) | 0.01(0.00,0.01) | 0.26(0.23,0.29) | 0.62(0.57,0.66) |
| Inattention_nested | 0.01(0.00,0.02) | 0.53(0.53,0.90) | 0.98(-0.99,1.00) | 0.33(0.29,0.37) | 0.35(0.27,0.43) | 0.73(0.70,0.76) | 0.37(0.33,0.41) | 0.00(0.00,0.00) | 0.00(0.00,0.00) | 0.67(0.63,0.71) | 0.15(0.08,0.22) | 0.27(0.24,0.30) | 0.62(0.58,0.66) |

Note: The best-fitting models are Cholesky decomposition models;

d_11_ represents the dominant genetic effect on hyperactivity-impulsivity/inattention; 2 on 3 represents the standardized causal effect from binge eating to BMI.

a_11_ represents the first genetic factor (A1) loading on hyperactivity-impulsivity/inattention, a_12_ represents the first genetic factor (A1) loading on binge eating, a_13_ represents the first genetic factor (A1) loading on BMI, a_22_ represents the second genetic factor (A2) loading on binge eating, a_23_ represents the second genetic factor (A2) loading on BMI, and a_33_ represents the third genetic factor (A3) loading on BMI; e_11_ represents the first non-shared environmental factor (E1) loading on hyperactivity-impulsivity/inattention, e_12_ represents the first non-shared environmental factor (E1) loading on binge eating, e13 represents the first non-shared environmental factor (E1) loading on BMI, e_22_ represents the second non-shared environmental factor (E2) loading on binge eating, e_23_ represents the second non-shared environmental factor (E2) loading on BMI, and e_33_ represents the third non-shared environmental factor (E3) loading on BMI (see Figure 2a).
